# COVID-19-Associated Hospitalizations Among Children Less Than 12 Years of Age in the United States

**DOI:** 10.1101/2022.01.04.22268742

**Authors:** Manuela Di Fusco, Shailja Vaghela, Mary M Moran, Jay Lin, Jessica E Atwell, Deepa Malhotra, Thomas Scassellati Sforzolini, Alejandro Cane, Jennifer L Nguyen, Leah J McGrath

**Author notes:** **Corresponding author** Manuela Di Fusco, Patient and Health Impact, Health Economics and Outcomes Research, Pfizer, Inc., New York, NY, USA.

## Abstract

**Objectives:** To describe the characteristics, healthcare resource use and costs associated with initial hospitalization and readmissions among pediatric patients with COVID-19 in the US.

**Methods:** Hospitalized pediatric patients, 0-11 years of age, with a primary or secondary discharge diagnosis code for COVID-19 (ICD-10 code U07.1) were selected from 1 April 2020 through 30 September 2021 in the US Premier Healthcare Database Special Release (PHD SR). Patient characteristics, hospital length of stay (LOS), in-hospital mortality, hospital costs, hospital charges, and COVID-19-associated readmission outcomes were evaluated and stratified by age groups (0-4, 5-11), four COVID-19 disease progression states based on intensive care unit (ICU) and invasive mechanical ventilation (IMV) usage, and three sequential calendar periods. Sensitivity analyses were performed using the US HealthVerity claims database and restricting the analyses to primary discharge code.

**Results:** Among 4,573 hospitalized pediatric patients aged 0-11 years, 68.0% were 0-4 years and 32.0% were 5-11 years, with a mean (median) age of 3.2 (1) years; 56.0% were male, and 67.2% were covered by Medicaid. Among the overall study population, 25.7% had immunocompromised condition(s), 23.1% were admitted to the ICU and 7.3% received IMV. The mean (median) hospital LOS was 4.3 (2) days, hospital costs and charges were $14,760 ($6,164) and $58,418 ($21,622), respectively; in-hospital mortality was 0.5%. LOS, costs, charges, and in-hospital mortality increased with ICU admission and/or IMV usage. In total, 2.1% had a COVID-19-associated readmission. Study outcomes appeared relatively more frequent and/or higher among those 5-11 than those 0-4. Results using the HealthVerity data source were generally consistent with main analyses.

**Limitations:** This retrospective administrative database analysis relied on coding accuracy and inpatient admissions with validated hospital costs.

**Conclusions:** These findings underscore that children aged 0-11 years can experience severe COVID-19 illness requiring hospitalization and substantial hospital resource use, further supporting recommendations for COVID-19 vaccination.

## Introduction

Although COVID-19-associated hospitalization rates have been reported to be lower in the pediatric population than in adults [1], COVID-19 can cause severe illness in children and adolescents and lead to serious long-term complications [2–6]. Prior studies conducted in the United States (US) have shown that 10-33% of children and adolescents with COVID-19-associated hospitalizations were admitted to an intensive care unit (ICU), up to 10% required invasive mechanical ventilation (IMV), and up to 2% died, depending on the time periods of assessment [2,3,6,7]. Two of these studies by the Centers for Disease Control and Prevention (CDC), with time periods that extended into when the SARS-CoV-2 delta variant was circulating in the US, found that COVID-19 hospitalizations among children and adolescents, 0-17 years of age, increased substantially in the latter weeks of July and August of 2021, especially among children 0-4 years of age [2,3]. Although important for understanding hospitalization patterns and disease severity in the pediatric population, these previous studies have been limited in terms of sample size, age group stratification, and/or geographic scale.

With the reopening of schools and the predominance of the delta variant from mid-July through December 2021 in the US [8], more detailed data on health outcomes and the hospital economic burden of pediatric COVID-19-associated hospitalizations among different pediatric age groups is warranted. Furthermore, readmission outcomes of pediatric populations with COVID-19-associated hospitalizations remain undescribed in the scientific literature. On October 29, 2021, the Pfizer-BioNTech COVID-19 vaccine received emergency use authorization for children 5-11 years of age in the US [9]. Further safety and efficacy data in children 6 months to 5 years of age from the phase 2/3 study [10] are forthcoming [11]. Specific data on health outcomes and the hospital economic burden of pediatric populations hospitalized with COVID-19 may be useful for decision makers as they plan policy implementations and future investments in COVID-19 prevention and treatments for children.

## Methods

### Study design and data sources

The methods of this study were similar to those from the previous study of Di Fusco et al. [7]. A retrospective cohort study was designed and main analyses conducted using data from the US Premier Healthcare Database Special Release (PHD SR) to address the study objective of characterizing hospitalization outcomes in the pediatric population less than 12 years of age. The PHD SR is a large, hospital-based, service-level, all-payer database representative of approximately 20% of all hospital admissions annually in the US [12]. Administrative healthcare data elements in the PHD SR are obtained from hospital discharge information and include patient demographic information, visit-level information, including hospital LOS and in-hospital mortality, clinical diagnoses, and medication information, in addition to hospital characteristics and hospital costs and charges. An additional sensitivity analysis was performed, and all analyses were repeated using claims data from the US HealthVerity Real Time Insights and Evidence (HV RTIE) database, a nationwide administrative healthcare claims data source. The key data elements contained within the HV database are relatively similar to that included in the PHD SR (e.g., patient demographic information, excluding race/ethnicity data, visit-level information, hospital characteristics and hospitalization payment/cost data). All data are de-identified and both databases are compliant with the Health Insurance Portability and Accountability Act (HIPAA) regulations.

### Study population

Pediatric patients aged 0-11 years hospitalized with COVID-19 were identified between 1 April 2020 and 30 September 2021. COVID-19 was identified using the International Classification of Diseases, Tenth Revision (ICD-10-CM) diagnosis code of U07.1 [13], in either the primary or secondary position. COVID-19 diagnosis was required to be designated as present on admission (POA), which identifies conditions as present at the time the inpatient admission occurred. The date of the first inpatient admission (index hospitalization) was designated as the index date.

### Demographics, hospitalization characteristics, and comorbid conditions

The demographic and hospitalization characteristics evaluated included age, sex, race/ethnicity, insurance type, hospital Census region, hospital population served (urban/rural), COVID-19 diagnosis position, calendar period of admission according to delta-relevant time periods (see measured outcomes section), calendar month of admission, and discharge status. Additionally, comorbid conditions were identified by using ICD-10-CM diagnosis and procedure codes, Current Procedural Terminology-4 (CPT-4) codes, and/or Healthcare Common Procedure Coding System (HCPCS) codes in primary or secondary positions on hospital records. While COVID-19 vaccination was not yet authorized for the pediatric study population 0-11 years of age during the time period of this study, evidence of COVID-19 vaccination was assessed based on CPT-4 codes and National Drug Codes [14] recorded on healthcare claims/hospital records as part of initial feasibility analyses.

In addition to stratification by age groups, 0-11, 0-4, and 5-11 years of age, demographic characteristics, hospitalization characteristics, and comorbid conditions, were stratified by four mutually exclusive COVID-19 disease progression states based on ICU admission and IMV usage as follows: 1) hospitalization without ICU admission or IMV usage (general ward); 2) hospitalization with ICU admission but without IMV usage (ICU but no IMV); 3) hospitalization without ICU admission but with IMV usage (no ICU but IMV), and 4) hospitalization with ICU admission and IMV usage (ICU and IMV) [7]. ICU admission and IMV usage were identified using revenue codes, HCPCS codes, and/or CPT-4 codes. IMV included extracorporeal membrane oxygenation (ECMO).

### Measured outcomes

The outcomes of interest included index hospitalization LOS, in-hospital mortality, costs and charges. While the primary focus of the cost analyses were hospital costs, hospital charges and a charge to cost ratio were also assessed. Hospital costs are reflective of the costs incurred by hospitals, while hospital charges represent standard list prices for hospital services and are generally higher than the actual cost of care incurred by hospitals. COVID-19-associated hospital readmissions were also evaluated with the associated outcomes of hospital LOS, in-hospital mortality, costs and charges. A readmission was defined as any first subsequent hospitalization with a COVID-19 diagnosis present as a primary or secondary diagnosis that occurred within two months (60 days) of the index COVID-19-associated hospitalization discharge date, similar to as in a prior CDC study [15]. An exploratory analysis was conducted to assess the proportion of readmissions occurring within 1 month (30 days) of the index COVID-19-associated hospitalization, and the proportion of COVID-19-associated readmissions out of all-cause readmissions.

All measured outcomes were stratified by age groups and the four COVID-19 disease progression states. Additionally, the study outcomes were stratified by the following time periods: pre-delta variant: April 2020-April 2021; emerging delta variant predominance: May 2021-June 2021; and during delta variant predominance: July 2021-September 2021 [8].

### Sensitivity analyses

A sensitivity analysis was conducted using the HV RTIE database to compare patterns with the main analyses using the PHD SR database. An additional sensitivity analysis was conducted among patients 0-11 years of age with an index hospitalization with a primary diagnosis of COVID-19 POA during the overall study period (1 April 2020-30 September 2021).

### Statistical analyses

Descriptive statistics summarized patient demographic and hospitalization characteristics, clinical characteristics, index hospitalization LOS, index hospitalization costs and charges, and index in-hospital mortality, in addition to the measured readmission outcomes. Counts and percentages were reported for categorical variables. Continuous variables were summarized using means, standard deviations (SD), medians, and first (Q1) and third (Q3) quartiles. All statistical analyses were carried out in SAS 9.4 (SAS Institute, Cary, NC).

### Conduct and ethics statements

This study followed the Strengthening the Reporting of Observational Studies in Epidemiology (STROBE) reporting guidelines [16]. This study was deemed exempt from Institutional Review Board (IRB) review pursuant to the terms of the US Department of Health and Human Service’s Policy for Protection of Human Research Subjects at 45 C.F.R. 46.104(d); category 4 exemption (Sterling IRB, Atlanta, GA, waived ethical approval for this work).

## Results

### Main analyses: PHD SR

#### Study population

The patient selection flow is shown in Figure 1. Of the total inpatient admissions reported in the PHD SR between 1 April 2020 and 30 September 2021, 15.0% (1,341,087) were children aged ≤11 years. Of those, 0.3% (N=4,573) had a COVID-19-associated hospitalization with a recorded primary or secondary diagnosis of COVID-19 and COVID-19 recorded as POA. Among these 4,573 pediatric patients, 68.0% were 0-4 years of age and the remaining 32.0% were 5-11 years of age.

**Figure 1.**
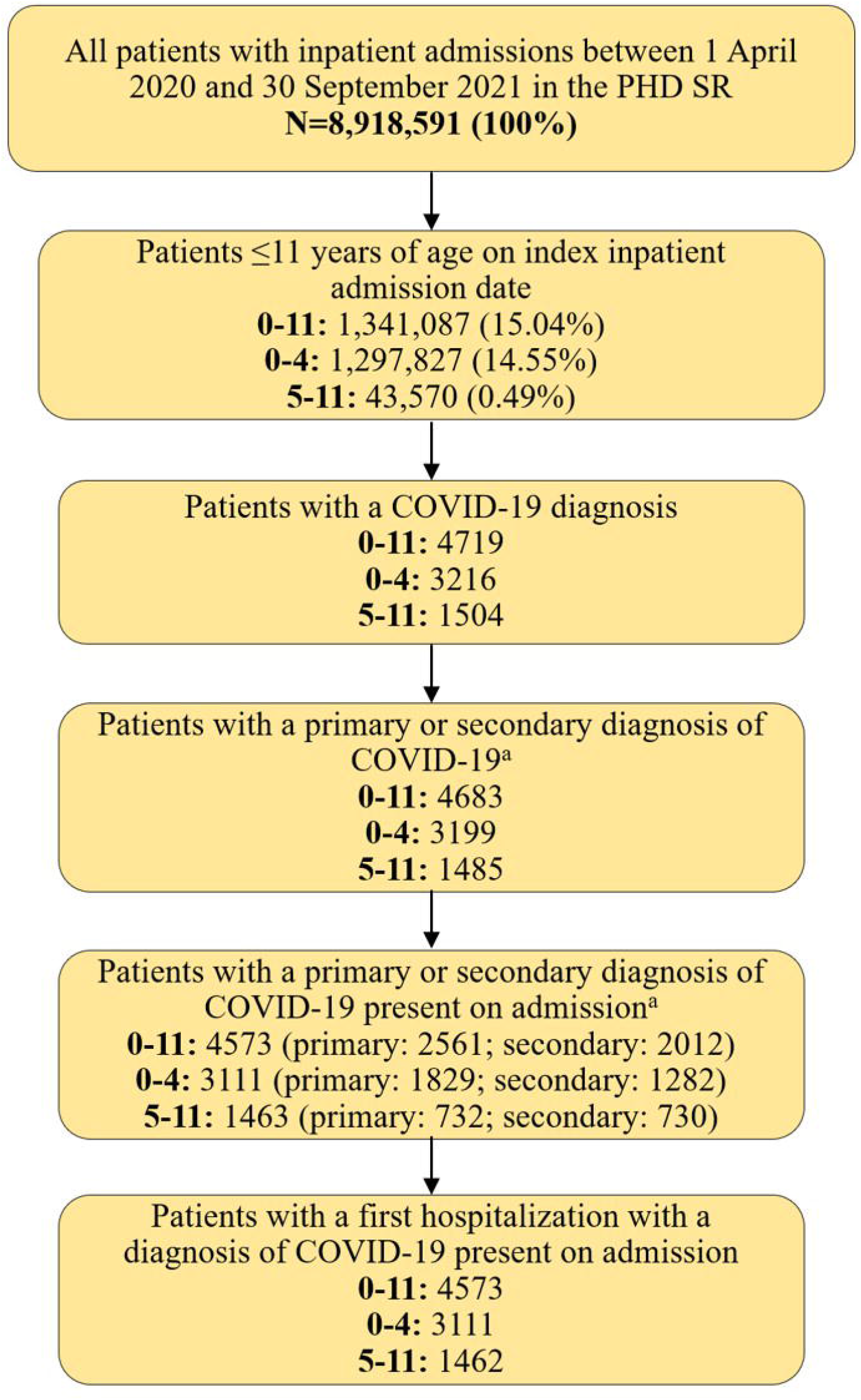
Flow of selection of study cohort ^a^ Patients may have had a COVID-19 diagnosis listed in both the primary and secondary positions, thus these categories may not sum to 100%.

#### Demographics, hospitalization characteristics, and comorbid conditions

Patient demographics, hospitalization characteristics and comorbid conditions of the pediatric population aged 0-11 years, 0-4 years, and 5-11 years, stratified by disease progression states based on ICU and IMV usage, are summarized in Table 1, supplementary Table 1, and supplementary Table 2, respectively.

**Table 1.** PHD SR: Demographics, hospitalization characteristics, and comorbid conditions among children aged 0-11 years.

|  | **Overall study population**<br>**N=4573 (100%)** |  | **Without ICU or IMV**<br>**N=3481 (76.1%)** |  | **With ICU, but without IMV**<br>**N=757 (16.6%)** |  | **Without ICU, but with IMV**<br>**N=34 (0.7%)** |  | **With ICU and IMV**<br>**N=301 (6.6%)** |  |
| --- | --- | --- | --- | --- | --- | --- | --- | --- | --- | --- |
| **Age (years)** |  |  |  |  |  |  |  |  |  |  |
| Median, Q1-Q3 | 1 | 0-6 | 1 | 0-6 | 4 | 1-8 | 3 | 0-6 | 2 | 0-6 |
| Mean, SD | 3.2 | 3.8 | 2.9 | 3.7 | 4.4 | 3.9 | 3.5 | 3.7 | 3.4 | 3.8 |
|  | *N* | % | *N* | % | *N* | % | *N* | % | *N* | % |
| **Age group** |  |  |  |  |  |  |  |  |  |  |
| 0-4 years | 3111 | 68.0 | 2472 | 71.0 | 410 | 54.2 | 24 | 70.6 | 205 | 68.1 |
| 5-11 years | 1462 | 32.0 | 1,009 | 29.0 | 347 | 45.8 | 10 | 29.4 | 96 | 31.9 |
| **Sex** |  |  |  |  |  |  |  |  |  |  |
| Male | 2560 | 56.0 | 1921 | 55.2 | 455 | 60.1 | 19 | 55.9 | 165 | 54.8 |
| Female | 2013 | 44.0 | 1560 | 44.8 | 302 | 39.9 | 15 | 44.1 | 136 | 45.2 |
| **Race/ethnicity** |  |  |  |  |  |  |  |  |  |  |
| White non-Hispanic | 1194 | 26.1 | 949 | 27.3 | 160 | 21.1 | 15 | 44.1 | 70 | 23.3 |
| Black non-Hispanic | 758 | 16.6 | 549 | 15.8 | 146 | 19.3 | 3 | 8.8 | 60 | 19.9 |
| Asian non-Hispanic | 106 | 2.3 | 85 | 2.4 | 12 | 1.6 | 1 | 2.9 | 8 | 2.7 |
| Other non-Hispanic | 256 | 5.6 | 188 | 5.4 | 39 | 5.2 | 5 | 14.7 | 24 | 8.0 |
| White Hispanic | 795 | 17.4 | 595 | 17.1 | 147 | 19.4 | 4 | 11.8 | 49 | 16.3 |
| Black Hispanic | 48 | 1.0 | 38 | 1.1 | 9 | 1.2 | 0 | 0.0 | 1 | 0.3 |
| Asian Hispanic | 2 | 0.0 | 2 | 0.1 | 0 | 0.0 | 0 | 0.0 | 0 | 0.0 |
| Other Hispanic | 541 | 11.8 | 417 | 12.0 | 86 | 11.4 | 3 | 8.8 | 35 | 11.6 |
| Unknown<sup>a</sup> | 873 | 19.1 | 658 | 18.9 | 158 | 20.9 | 3 | 8.8 | 54 | 17.9 |
| **Insurance type** |  |  |  |  |  |  |  |  |  |  |
| Commercial | 1086 | 23.7 | 860 | 24.7 | 163 | 21.5 | 6 | 17.6 | 57 | 18.9 |
| Medicaid | 3072 | 67.2 | 2311 | 66.4 | 521 | 68.8 | 26 | 76.5 | 214 | 71.1 |
| Medicare | 15 | 0.3 | 12 | 0.3 | 2 | 0.3 | 0 | 0.0 | 1 | 0.3 |
| Other | 301 | 6.6 | 230 | 6.6 | 51 | 6.7 | 1 | 2.9 | 19 | 6.3 |
| Uninsured | 99 | 2.2 | 68 | 2.0 | 20 | 2.6 | 1 | 2.9 | 10 | 3.3 |
| **Hospital Census region** |  |  |  |  |  |  |  |  |  |  |
| Northeast | 668 | 14.6 | 526 | 15.1 | 89 | 11.8 | 4 | 11.8 | 49 | 16.3 |
| Midwest | 598 | 13.1 | 464 | 13.3 | 91 | 12.0 | 6 | 17.6 | 37 | 12.3 |
| South | 2511 | 54.9 | 1,865 | 53.6 | 468 | 61.8 | 17 | 50.0 | 161 | 53.5 |
| West | 796 | 17.4 | 626 | 18.0 | 109 | 14.4 | 7 | 20.6 | 54 | 17.9 |
| ***Hospital population served*** |  |  |  |  |  |  |  |  |  |  |
| Urban | 4294 | 93.9 | 3304 | 94.9 | 675 | 89.2 | 28 | 82.4 | 287 | 95.3 |
| Rural | 279 | 6.1 | 177 | 5.1 | 82 | 10.8 | 6 | 17.6 | 14 | 4.7 |
| ***COVID-19 diagnosis position*** |  |  |  |  |  |  |  |  |  |  |
| Primary | 2561 | 56.0 | 1,994 | 57.3 | 416 | 55.0 | 17 | 50.0 | 134 | 44.5 |
| Secondary | 2012 | 44.0 | 1,487 | 42.7 | 341 | 45.0 | 17 | 50.0 | 167 | 55.5 |
| ***Calendar period of admission*** |  |  |  |  |  |  |  |  |  |  |
| April 2020-April 2021 | 2934 | 64.2 | 2,208 | 63.4 | 518 | 68.4 | 18 | 52.9 | 190 | 63.1 |
| May 2021-June 2021 | 233 | 5.1 | 177 | 5.1 | 38 | 5.0 | 1 | 2.9 | 17 | 5.6 |
| July 2021-September 2021 | 1,406 | 30.7 | 1,096 | 31.5 | 201 | 26.6 | 15 | 44.1 | 94 | 31.2 |
| ***Discharge status<sup>b</sup>*** |  |  |  |  |  |  |  |  |  |  |
| Home/self-care | 4298 | 94.0 | 3,339 | 95.9 | 719 | 95.0 | 22 | 64.7 | 218 | 72.4 |
| Expired/died | 23 | 0.5 | 0 | 0.0 | 1 | 0.1 | 1 | 2.9 | 21 | 7.0 |
| Transferred to other facility | 144 | 3.1 | 97 | 2.8 | 20 | 2.6 | 7 | 20.6 | 20 | 6.6 |
| ***Comorbid conditions*** |  |  |  |  |  |  |  |  |  |  |
| Number of comorbidities: 0 | 2563 | 56.1 | 2166 | 62.2 | 277 | 36.6 | 18 | 52.9 | 102 | 33.9 |
| Number of comorbidities: 1-2 | 1792 | 39.2 | 1210 | 34.8 | 422 | 55.8 | 13 | 38.2 | 147 | 48.8 |
| Number of comorbidities: $\geq 3$ | 218 | 4.8 | 105 | 3.0 | 58 | 7.7 | 3 | 8.8 | 52 | 17.3 |
| Median, Q1-Q3 | 0 | 0-1 | 0 | 0-1 | 1 | 0-2 | 0 | 0-1 | 1 | 0-2 |
| Mean, SD | 0.7 | 0.9 | 0.5 | 0.8 | 1.0 | 1.0 | 0.8 | 1.0 | 1.3 | 1.3 |
| ***Comorbid conditions*** |  |  |  |  |  |  |  |  |  |  |
| Diabetes | 83 | 1.8 | 30 | 0.9 | 50 | 6.6 | 0 | 0.0 | 3 | 1.0 |
| Obesity/overweight | 190 | 4.2 | 110 | 3.2 | 53 | 7.0 | 3 | 8.8 | 24 | 8.0 |
| Hypertension | 108 | 2.4 | 49 | 1.4 | 28 | 3.7 | 0 | 0.0 | 31 | 10.3 |
| CKD or ESRD | 241 | 5.3 | 100 | 2.9 | 78 | 10.3 | 1 | 2.9 | 62 | 20.6 |
| Neurological disease | 351 | 7.7 | 199 | 5.7 | 69 | 9.1 | 8 | 23.5 | 75 | 24.9 |
| Psychiatric disorders | 99 | 2.2 | 71 | 2.0 | 17 | 2.2 | 1 | 2.9 | 10 | 3.3 |
| Malignancy (solid or hematologic) | 178 | 3.9 | 142 | 4.1 | 28 | 3.7 | 0 | 0.0 | 8 | 2.7 |
| Asthma/reactive airway disease | 473 | 10.3 | 315 | 9.0 | 115 | 15.2 | 2 | 5.9 | 41 | 13.6 |
| Chronic liver disease | 1 | 0.0 | 0 | 0.0 | 1 | 0.1 | 0 | 0.0 | 0 | 0.0 |
| Congenital heart condition | 29 | 0.6 | 15 | 0.4 | 7 | 0.9 | 0 | 0.0 | 7 | 2.3 |
| Congenital lung condition | 8 | 0.2 | 4 | 0.1 | 2 | 0.3 | 1 | 2.9 | 1 | 0.3 |
| Down syndrome/chromosomal anomaly | 74 | 1.6 | 45 | 1.3 | 13 | 1.7 | 3 | 8.8 | 13 | 4.3 |
| Immunocompromised<sup>c</sup> | 1173 | 25.7 | 762 | 21.9 | 292 | 38.6 | 5 | 14.7 | 114 | 37.9 |
| Autoimmune disease | 86 | 1.9 | 35 | 1.0 | 50 | 6.6 | 0 | 0.0 | 1 | 0.3 |
| Sickle cell | 117 | 2.6 | 88 | 2.5 | 21 | 2.8 | 0 | 0.0 | 8 | 2.7 |
| Disability<sup>d</sup> | 56 | 1.2 | 36 | 1.0 | 8 | 1.1 | 2 | 5.9 | 10 | 3.3 |
| Transplant (bone marrow and organ) | 33 | 0.7 | 25 | 0.7 | 4 | 0.5 | 1 | 2.9 | 3 | 1.0 |

<sup>a</sup> Unknown refers to either one of, or both, race and ethnicity are unknown.
<sup>b</sup> Reported only top three discharge status here, remaining such as skilled nursing facility, home health organization, hospice, rehabilitation facility, and/or other are not listed.
<sup>c</sup> Immunocompromised conditions included HIV/AIDS, malignancy, transplants, rheumatologic/other inflammatory conditions, primary immunodeficiency, CKD/ESRD, and other immune conditions.
<sup>d</sup> Includes neurologic, neurodevelopmental, intellectual, physical, vision or hearing impairment.
AIDS: Acquired immunodeficiency syndrome. CKD: Chronic kidney disease; ESRD: End stage renal disease; HIV: Human immunodeficiency virus; ICU: Intensive care unit; IMV: Invasive mechanical ventilation; PHD SR: Premier Healthcare Database Special Release; Q1: First quartile; Q3: Third quartile; SD: Standard deviation.

The mean (median) age of the overall pediatric study population aged 0-11 years, was 3.2 (1) years, and was 0.9 (0) years in the age group of 0-4 years and 8.3 (8) years in the age group of 5-11 years. Among the 0-11 year cohort, 56.0% were male, 26.1% were White non-Hispanic, 30.3% were Hispanic, 16.6% were Black non-Hispanic, and 19.1% had unknown race/ethnicity. Over two-thirds (67.2%) were insured by Medicaid and 23.7% had commercial insurance coverage. Most COVID-19-associated hospitalizations occurred in urban hospitals (93.9%). Similar demographic distributions were observed in both 0-4 and 5-11 age groups.

In the overall study population aged 0-11 years, 56.1% had no comorbidities, 39.2% had one or two comorbidities, and 4.8% children had ≥3 comorbid conditions. The mean number of comorbidities was 0.7, which increased with disease progression from 0.5 in the general ward to 1.3 in the ICU and IMV cohort. Additionally, approximately one-quarter (25.7%) had one or more immunocompromised conditions; rheumatological/other inflammatory conditions, primary immunodeficiencies, and chronic kidney disease (CKD)/end stage renal disease (ESRD) were the most prevalent. Other prevalent comorbid conditions included asthma/reactive airway disease (10.3%) and neurological disease (7.7%). Immunocompromised conditions were even more prevalent (>35%) among those with an ICU admission; CKD/ESRD and rheumatological conditions were reported most in both the ICU but no IMV and the ICU with IMV cohorts. This trend was generally true for neurologic disease and asthma/reactive airway disease.

Only 30.8% of the pediatric patients aged 0-4 years had any comorbidities, whereas 72.0% of the 5-11 years had one or more comorbidities with a mean number of comorbidities ranging from 1.0 in the general ward to 2.1 in the ICU and IMV. Furthermore, the prevalence of immunocompromised conditions was almost doubled in the 5-11 years age group (38.9%) compared to that in the age group of 0-4 years (19.4%).

#### Demographics, hospitalization characteristics and comorbidities at readmission

There were 96 pediatric cases (2.1%) that were readmitted to the hospital for COVID-19 within 2 months (60 days) (Table 2). Among those readmitted, 61.5% were 0-4 years and 38.5% were 5-11 years. The mean (median) age of the overall readmitted pediatric cases was 4.1 (3) years; it was 1.3 (1) years in the age group of 0-4 years and 8.5 (9) years in the age group of 5-11 years.

**Table 2.** PHD SR: Health outcomes and costs among children aged 0-11 years.

|  | Index hospitalizations | Hospital LOS, days |  | In-hospital mortality | Hospital costs<sup>a</sup> |  | Hospital charges |  |
| --- | --- | --- | --- | --- | --- | --- | --- | --- |
|  | N (%) | Median (Q1-Q3) | Mean (SD) | N (%) | Median (Q1-Q3) | Mean (SD) | Median (Q1-Q3) | Mean (SD) |
| **Index hospitalization status** |  |  |  |  |  |  |  |  |
| **All inpatient admissions** | 4573 (100) | 2 (1-4) | 4.3 (7.6) | 23 (0.5) | \$6164 (\$3432-\$14,095) | \$14,760 (\$30,447) | \$21,622 (\$10,885-\$49,602) | \$58,418 (\$163,278) |
| *Without ICU or IMV* | 3481 (76.1) | 2 (1-3) | 2.9 (3.4) | 0 (0) | \$4913 (\$3026-\$9130) | \$7888 (\$9026) | \$16,952 (\$9354-\$32,396) | \$28,669 (\$37,586) |
| *With ICU, but without IMV* | 757 (16.6) | 4 (2-6) | 5.3 (5.1) | 1 (0.1) | \$14,682 (\$6967-\$26,884) | \$21,704 (\$24,390) | \$52,225 (\$23,799-\$112,641) | \$85,202 (\$94,666) |
| *Without ICU, but with IMV* | 34 (0.7) | 3 (1-5) | 7.4 (16.1) | 1 (2.9) | \$9555 (\$5620-\$15,837) | \$25,770 (\$54,416) | \$33,684 (\$14,851-\$71,167) | \$96,747 (\$188,190) |
| *With ICU and IMV* | 301 (6.6) | 10 (5-21) | 17.0 (21.6) | 21 (7.0) | \$43,279 (\$23,495-\$91,662) | \$75,389 (\$84,372) | \$178,398 (\$88,325-\$404,648) | \$330,778 (\$526,076) |
| **Readmissions** |  |  |  |  |  |  |  |  |
| **Index hospitalization status** |  |  |  |  |  |  |  |  |
| **All inpatient admissions** | 96 (2.1) | 3 (2-5) | 5.0 (6.8) | 1 (1.0) | \$7965 (\$3816-\$17,653) | \$15,652 (\$24,112) | \$33,255 (\$11,617-\$75,017) | \$56,563 (\$72,020) |
| *Without ICU or IMV* | 74 (2.1) | 3 (2-4) | 4.6 (6.7) | 0 (0) | \$6811 (\$3505-\$10,760) | \$12,469 (\$21,407) | \$27,282 (\$11,077-\$55,325) | \$50,108 (\$65,677) |
| *With ICU, but without IMV* | 16 (2.1) | 3 (2-7.5) | 5.1 (4.5) | 1 (6.3) | \$12,234 (\$4071-\$23,383) | \$19,443 (\$24,594) | \$36,713 (\$11,036-\$73,571) | \$50,059 (\$51,910) |
| *Without ICU, but with IMV* | 1 (2.9) | 4 - | 4.0 - | 0 (0) | \$15,614 - | \$15,614 - | \$92,917 - | \$92,917 - |
| *With ICU and IMV* | 5 (1.7) | 4 (3-23) | 11.6 (12.4) | 0 (0) | \$20,911 (\$10,983-\$75,091) | \$42,482 (\$40,564) | \$123,614 (\$80,954-\$242,950) | \$165,632 (\$134,993) |

<sup>a</sup> Among patients with validated costs >\$0 (N=3,782 for index hospitalizations; N=81 for readmissions)
<sup>b</sup> Percentage among those with COVID-19-associated index hospitalizations, overall and stratified by the 4 COVID-19 disease progression states.
ICU: Intensive care unit; IMV: Invasive mechanical ventilation; LOS: Length of stay; PHD SR: Premier Healthcare Database Special Release; Q1: First quartile; Q3: Third quartile; SD: Standard deviation

Immunocompromised conditions were more prevalent among readmitted children compared to among those with index hospitalizations; 47.9% among those 0-11 years, 42.4% among those 0-4 years, and 56.8% among those 5-11 years. Approximately 85% (82 of 96) of all the COVID-19-associated readmissions occurred within 1 month (30 days); the 96 COVID-19-associated readmissions represented approximately one-third (32.3%) of the all-cause readmissions in the pediatric cohort (data available upon request).

#### Health outcomes and costs among children aged 0-11 years

The index hospitalization and readmission outcomes of the overall study population aged 0-11 years are shown in Table 2. Most pediatric patients (76.1%) were admitted to the general ward without ICU or IMV usage, 16.6% were admitted to the ICU but did not receive IMV, 0.7% received IMV but were not admitted to the ICU, and 6.6% were admitted to the ICU and received IMV. Overall, 23.1% were admitted to the ICU and 7.3% received IMV.

The mean (median) hospital LOS was 4.3 (2) days, which increased from 2.9 (2) days among those in the general ward to 17.0 (10) days among those admitted to the ICU with IMV usage. Similarly, in-hospital mortality (N=23, 0.5% of index hospitalization) increased from none in the general ward to 7.0% among those with ICU admission and IMV usage. The mean (median) costs and charges were $14,760 ($6,164) and $58,418 ($21,622), respectively. Non-zero hospital costs were recorded for 82.7% (N=3,782) of the pediatric study population. Based upon the mean (median) costs and charges reported, among those with validated hospital cost data, the mean (median) charge to cost ratio was calculated as 4.0 (3.5). The hospital costs and charges also increased with disease progression; for instance, the median cost in the general ward was $4,913, while among those admitted to the ICU with IMV usage, it was and $43,279.

Among the children with a COVID-19-associated hospital readmission within two months, the mean (median) hospital LOS was 5.0 (3) days, and costs and charges were $15,651 ($7,965) and $56,563 ($33,255), respectively. Thus, median hospital readmission LOS and median costs and charges were slightly higher than for index COVID-19-associated hospitalizations; however, since the readmission sample sizes were small, these trends could not be confirmed across the disease progression states of index hospitalizations. The frequency of readmission did not display a clear trend with COVID-19 illness severity during index hospitalizations; it ranged from 1.7% among those with an index hospitalization with both ICU admission and IMV usage to 2.9% among those without ICU admission but with IMV usage. Only 1 death was observed, corresponding to an in-hospital readmission mortality of ∼1%; the death was reported in the ICU no IMV cohort.

#### Health outcomes and costs among children aged 0-4 years and 5-11 years

The index hospitalization and readmission outcomes for the pediatric population aged 0-4 years and 5-11 years are shown in Table 3 and Table 4, respectively. Similar trends were observed in both age groups for IMV usage during hospitalization as observed in the overall pediatric population aged 0-11 years, however, 30.3% (N=443) of pediatric patients aged 5-11 years were admitted to an ICU compared to 19.8% (N=615) of those aged 0-4 years.

**Table 3.** PHD SR: Health outcomes and costs among children aged 0-4 years.

|  | Index hospitalizations | Hospital LOS, days |  | In-hospital mortality | Hospital costs<sup>a</sup> |  | Hospital charges |  |
| --- | --- | --- | --- | --- | --- | --- | --- | --- |
|  | N<br>(%) | Median<br>(Q1-Q3) | Mean<br>(SD) | N<br>(%) | Median<br>(Q1-Q3) | Mean<br>(SD) | Median<br>(Q1-Q3) | Mean<br>(SD) |
| **All inpatient admissions** | 3111<br>(100) | 2<br>(1-4) | 4.1<br>(8.1) | 14<br>(0.5) | \$5157<br>(\$3131-\$10,941) | \$12,379<br>(\$27,941) | \$17,403<br>(\$9671-\$37,896) | \$49,544<br>(\$169,375) |
| *Without ICU or IMV* | 2472<br>(79.5) | 2<br>(1-3) | 2.6<br>(2.5) | 0<br>(0) | \$4379<br>(\$2756-\$7540) | \$6653<br>(\$7429) | \$14,982<br>(\$8418-\$26,914) | \$23,717<br>(\$32,935) |
| *With ICU, but without IMV* | 410<br>(13.2) | 3<br>(2-6) | 5.2<br>(5.9) | 0<br>(0) | \$11,294<br>(\$5518-\$22,768) | \$18,446<br>(\$25,141) | \$38,975<br>(\$18,743-\$85,922) | \$68,255<br>(\$89,314) |
| *Without ICU, but with IMV* | 24<br>(0.8) | 3<br>(1.5-6) | 8.8<br>(18.9) | 0<br>(0) | \$8011<br>(\$4305-\$14,760) | \$25,546<br>(\$61,225) | \$21,666<br>(\$11,442-\$65,812) | \$88,601<br>(\$202,566) |
| *With ICU and IMV* | 205<br>(6.6) | 11<br>(4-23) | 18.3<br>(24.0) | 14<br>(6.8) | \$35,536<br>(\$17,613-\$89,591) | \$68,473<br>(\$78,343) | \$166,814<br>(\$63,734-\$357,483) | \$318,975<br>(\$567,167) |
|  | Readmissions | Hospital LOS, days |  | In-hospital mortality | Hospital costs<sup>a</sup> |  | Hospital charges |  |
|  | N<br>(%)<sup>b</sup> | Median<br>(Q1-Q3) | Mean<br>(SD) | N<br>(%) | Median<br>(Q1-Q3) | Mean<br>(SD) | Median<br>(Q1-Q3) | Mean<br>(SD) |
| **All inpatient admissions** | 59<br>(1.9) | 3<br>(2-5) | 5.4<br>(7.6) | 0<br>(0) | \$8260<br>(\$3486-\$18,483) | \$18,047<br>(\$29,785) | \$35,133<br>(\$12,241-\$87,309) | \$66,684<br>(\$86,544) |
| *Without ICU or IMV* | 48<br>(1.9) | 3<br>(1.5-5) | 4.9<br>(7.2) | 0<br>(0) | \$4908<br>(\$2961-\$12,328) | \$13,574<br>(\$25,849) | \$27,914<br>(\$10,855-\$75,017) | \$56,918<br>(\$77,371) |
| *With ICU, but without IMV* | 5<br>(1.2) | 3<br>(2-3) | 4.2<br>(4.4) | 0<br>(0) | \$9012<br>(\$4869-\$54,087) | \$29,478<br>(\$44,413) | \$23,220<br>(\$12,803-\$35,133) | \$56,241<br>(\$79,687) |
| *Without ICU, but with IMV* | 1<br>(4.2) | 4 | 4 | 0<br>(0) | \$15,614 | \$15,614 | \$92,917 | \$92,917 |
| *With ICU and IMV* | 5<br>(2.4) | 4<br>(3-23) | 11.6<br>(12.4) | 0<br>(0) | \$20,911<br>(\$10,983-\$75,091) | \$42,482<br>(\$40,564) | \$123,614<br>(\$80,954-\$242,950) | \$165,632<br>(\$134,993) |

<sup>a</sup> Among patients with validated costs >\$0 (N=2,574 for index hospitalizations; N=47 for readmissions).
<sup>b</sup> Percentage among those with COVID-19-associated index hospitalizations, overall and stratified by the 4 COVID-19 disease progression states.
ICU: Intensive care unit; IMV: Invasive mechanical ventilation; LOS: Length of stay; PHD SR: Premier Healthcare Database Special Release; Q1: First quartile; Q3: Third quartile; SD: Standard deviation

**Table 4.** PHD SR: Health outcomes and costs among children aged 5-11 years.

|  | Index hospitalizations | Hospital LOS, days |  | In-hospital mortality | Hospital costs<sup>a</sup> |  | Hospital charges |  |
| --- | --- | --- | --- | --- | --- | --- | --- | --- |
|  | N<br>(%) | Median<br>(Q1-Q3) | Mean<br>(SD) | N<br>(%) | Median<br>(Q1-Q3) | Mean<br>(SD) | Median<br>(Q1-Q3) | Mean<br>(SD) |
| **All inpatient admissions** | 1462<br>(100) | 3<br>(2-6) | 4.7<br>(6.6) | 9<br>(0.6) | \$9729<br>(\$4765-\$21,203) | \$19,835<br>(\$34,669) | \$34,227<br>(\$16,799-\$83,631) | \$77,304<br>(\$147,771) |
| *Without ICU or IMV* | 1009<br>(69.0) | 2<br>(1-4) | 3.5<br>(4.9) | 0<br>(0) | \$7075<br>(\$3755-\$13,439) | \$10,915<br>(\$11,538) | \$25,700<br>(\$12,716-\$49,342) | \$40,800<br>(\$44,841) |
| *With ICU, but without IMV* | 347<br>(23.7) | 5<br>(3-7) | 5.4<br>(4.1) | 1<br>(0.3) | \$19,653<br>(\$11,202-\$33,921) | \$25,727<br>(\$22,838) | \$68,604<br>(\$32,242-\$148,594) | \$105,226<br>(\$97,000) |
| *Without ICU, but with IMV* | 10<br>(0.7) | 3<br>(1-5) | 4.1<br>(3.9) | 1<br>(10.0) | \$12,207<br>(\$8707-\$25,759) | \$26,332<br>(\$35,301) | \$40,550<br>(\$31,869-\$183,814) | \$116,300<br>(\$156,185) |
| *With ICU and IMV* | 96<br>(6.6) | 9<br>(6-16) | 14.3<br>(15.3) | 7<br>(7.3) | \$56,578<br>(\$35,916-\$93,575) | \$89,392<br>(\$94,391) | \$195,697<br>(\$113,701-\$465,686) | \$355,984<br>(\$426,922) |
|  | Readmissions | Hospital LOS, days |  | In-hospital mortality | Hospital costs<sup>a</sup> |  | Hospital charges |  |
|  | N<br>(%)<sup>b</sup> | Median<br>(Q1-Q3) | Mean<br>(SD) | N<br>(%) | Median<br>(Q1-Q3) | Mean<br>(SD) | Median<br>(Q1-Q3) | Mean<br>(SD) |
| **All inpatient admissions** | 37<br>(2.5) | 3<br>(2-5) | 4.5<br>(5.4) | 1<br>(2.7) | \$7780<br>(\$4544-\$17,653) | \$12,341<br>(\$12,382) | \$28,145<br>(\$9874-\$52,692) | \$40,424<br>(\$34,426) |
| *Without ICU or IMV* | 26<br>(2.6) | 3<br>(2-3) | 4.1<br>(5.7) | 0<br>(0) | \$6965<br>(\$4544-\$10,631) | \$10,690<br>(\$11,410) | \$27,282<br>(\$11,077-\$52,218) | \$37,536<br>(\$32,982) |
| *With ICU, but without IMV* | 11<br>(3.2) | 5<br>(2-8) | 5.5<br>(4.7) | 1<br>(9.1) | \$12,234<br>(\$4071-\$23,383) | \$15,793<br>(\$14,147) | \$45,034<br>(\$9533-\$93,515) | \$47,249<br>(\$38,394) |
| *Without ICU, but with IMV* | 0<br>(0) | - | - | 0<br>(0) | - | - | - | - |
| *With ICU and IMV* | 0<br>(0) | - | - | 0<br>(0) | - | - | - | - |

<sup>a</sup> Among patients with validated costs >\$0 (N=1,208 for index hospitalizations; N=34 for readmissions).
<sup>b</sup> Percentage among those with COVID-19-associated index hospitalizations, overall and stratified by the 4 COVID-19 disease progression states.
ICU: Intensive care unit; IMV: Invasive mechanical ventilation; LOS: Length of stay; PHD SR: Premier Healthcare Database Special Release; Q1: First quartile; Q3: Third quartile; SD: Standard deviation

The mean (median) hospital LOS was longer among those aged 5-11 years at 4.7 (3) days compared to 4.1 (2) days among those aged 0-4 years. Most in-hospital mortality was reported in the ICU and IMV cohort for both age groups; mortality rates were slightly higher among children 5-11 years of age (7.3%) than among those 0-4 years of age (6.8%) in this disease progression state. The mean (median) hospitalization cost in the age group of 0-4 years was $12,378 ($5,157), while it was higher in the age group of 5-11 years at $19,835 ($9,729). Similarly, mean (median) charges were also higher for those 5-11 years at $77,303 ($34,226) compared to those 0-4 years at $49,543 ($17,402). Hospital LOS, costs, and charges increased with disease progression state in both age groups.

Among the pediatric patients aged 0-4 years with index COVID-19-associated hospitalizations, 1.9% (N=59) had a COVID-19-associated hospital readmission; the mean (median) hospital LOS was 5.4 (3) days, and hospital costs and charges were $18,047 ($8,260) and $66,684 ($35,133), respectively. The frequency of readmission ranged from 1.2% among those in the index admission ICU but no IMV group to 4.2% in the no ICU but IMV cohort. Among those aged 5-11 years, 2.5% (N=37) had a COVID-19-associated hospital readmission; the mean (median) hospital LOS was 4.5 (3) days, and hospital costs and charges were $12,341 ($7,780) and $40,423 ($28,145), respectively. No deaths occurred among patients aged 0-4 years with a COVID-19-associated readmission; whereas 1 death occurred in the ICU but no IMV cohort among patients aged 5-11 years.

#### Index hospitalization and readmission outcomes by calendar period among children aged 0-11 years

The index hospitalization and readmission outcomes in the overall study population aged 0-11 years stratified by COVID-19 disease progression states and the three calendar periods are shown in Table 5 and Table 6, respectively. Of all the pediatric COVID-19-associated inpatient admissions, 64.2% occurred prior to the emergence of the delta variant in the US (April 2020-April 2021 [pre-delta]), 5.1% occurred when the delta variant began circulating (May 2021-June 2021 [transition]), and 30.7% occurred during delta variant predominance (July 2021-September 2021 [delta]) (Table 1). By calendar month, the frequency of pediatric COVID-19-associated hospital admissions gradually increased from 6.7% in November 2020, to 8.7% in December 2020, to 9.8% in January 2021, then declined until June 2021, and then increased again in the months of delta predominance: July 2021 (5.6%), August 2021 (13.6%), and September 2021(11.5%) (Figure 2). Across the three study periods, the frequency of ICU admission (pre-delta: 24.1%; transition: 23.6%; delta: 21.0%) and IMV usage (pre-delta: 7.1%; transition: 7.7%; delta: 7.8%) was similar. Additionally, the proportions of the pediatric population with both ICU admission and IMV usage and the proportions without ICU admission or IMV usage were generally similar across the study periods. The frequency of a COVID-19-associated readmission within two months was 2.4% in the pre-delta period, 1.7% in the transition period, and 1.6% during delta period.

**Table 5.**
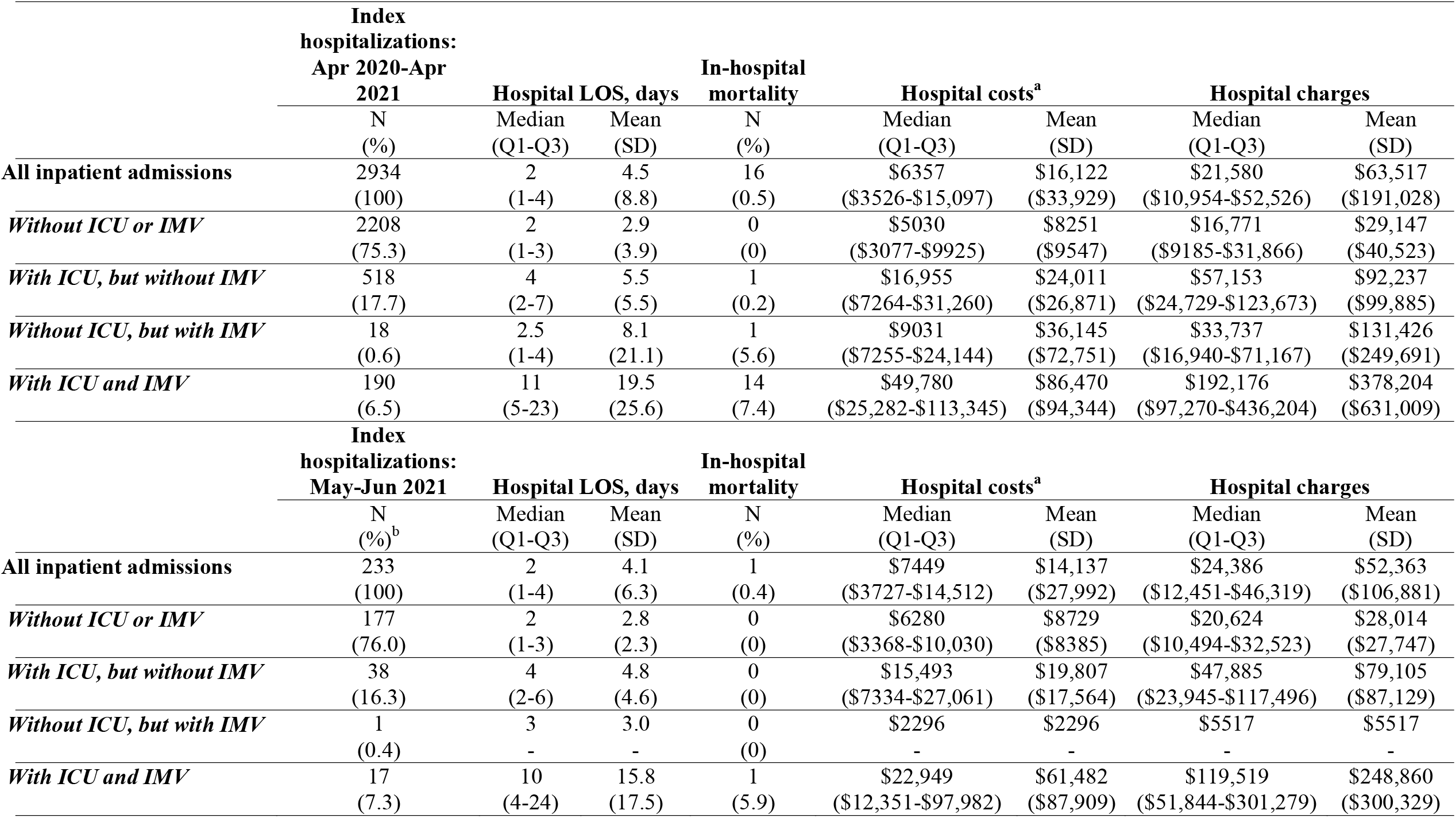
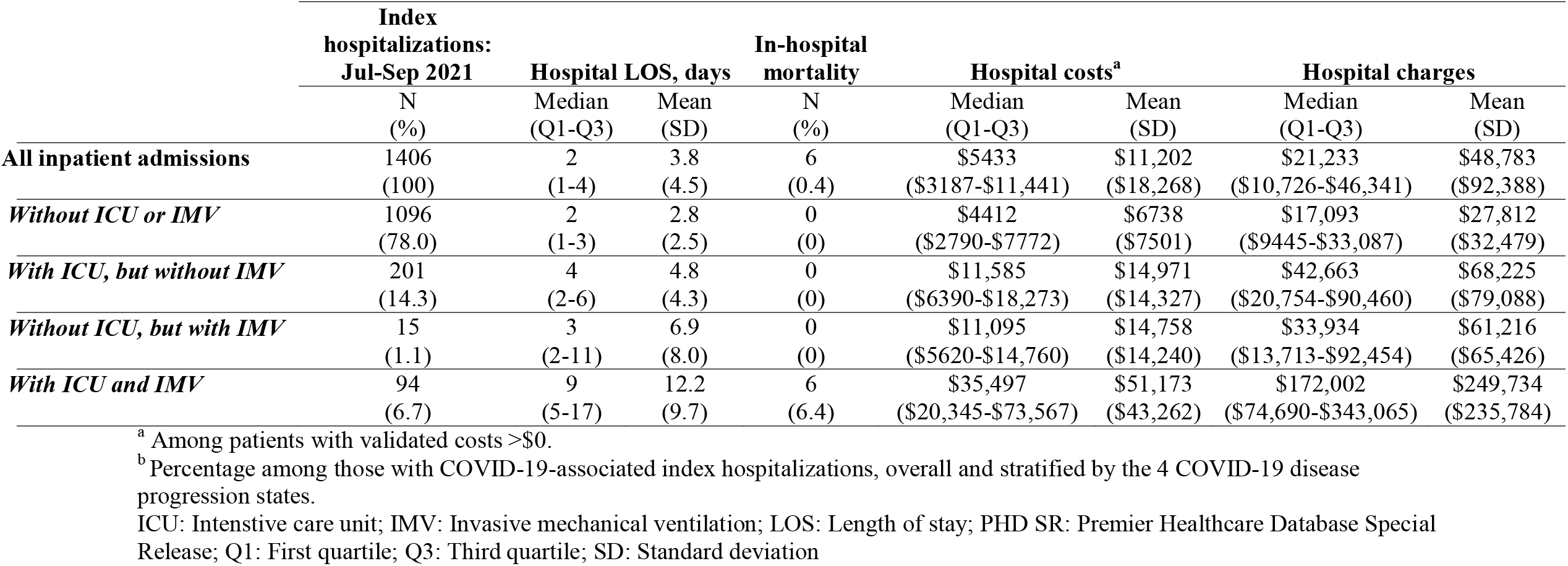
PHD SR: Index hospitalization outcomes by calendar period among children aged 0-11 years.

**Table 6.**
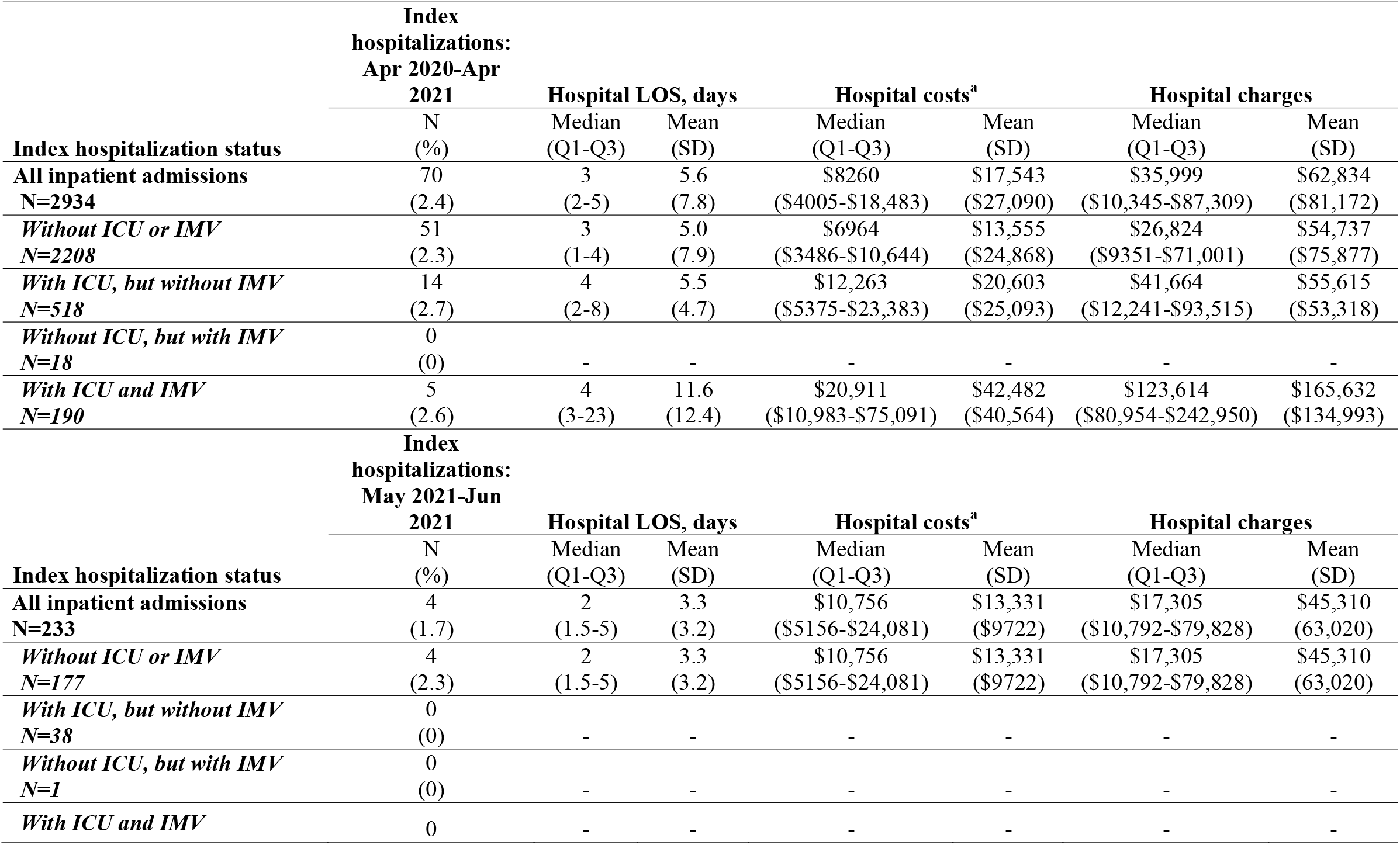
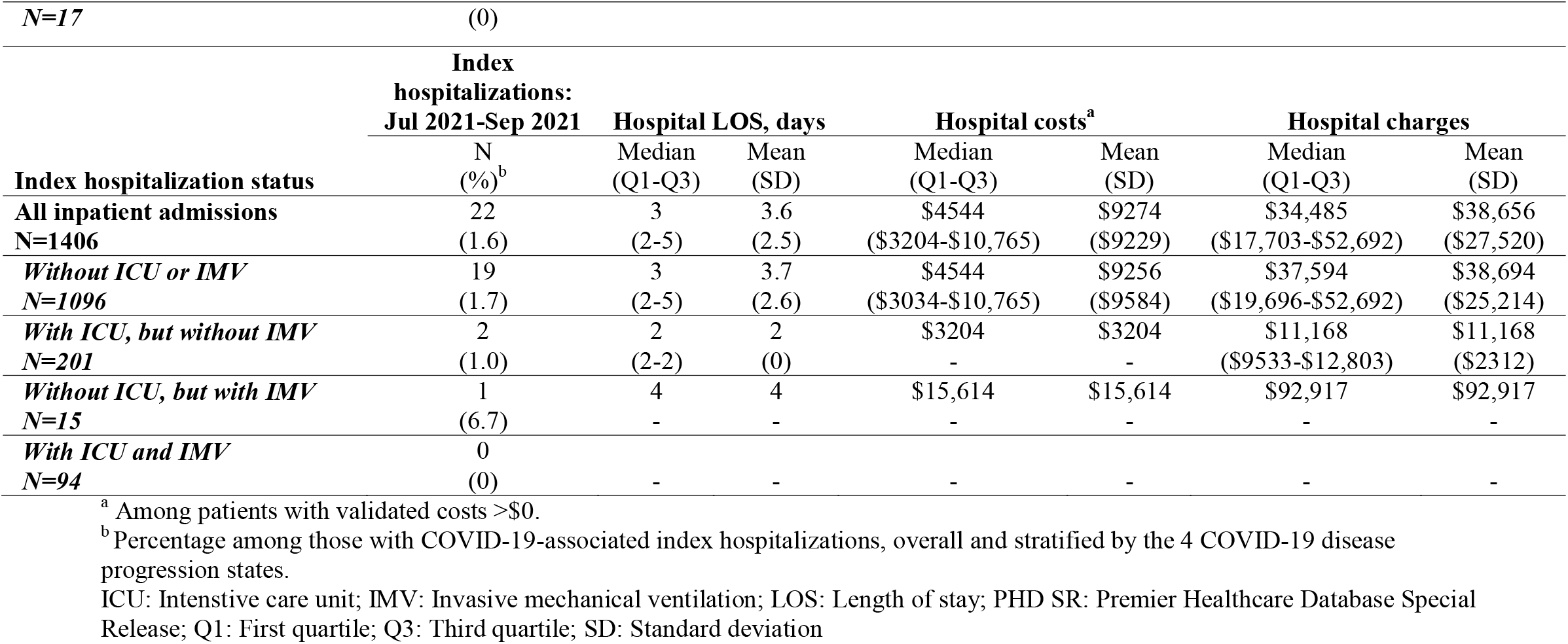
PHD SR: Readmission outcomes by calendar period among children aged 0-11 years.

|  | Index hospitalizations:<br>Apr 2020-Apr 2021 |  |  |  |  |  |  |
| --- | --- | --- | --- | --- | --- | --- | --- |
|  | N | Hospital LOS, days |  | Hospital costs<sup>a</sup> |  | Hospital charges |  |
| Index hospitalization status | (%) | Median (Q1-Q3) | Mean (SD) | Median (Q1-Q3) | Mean (SD) | Median (Q1-Q3) | Mean (SD) |
| **All inpatient admissions**<br>**N=2934** | 70<br>(2.4) | 3<br>(2-5) | 5.6<br>(7.8) | \$8260<br>(\$4005-\$18,483) | \$17,543<br>(\$27,090) | \$35,999<br>(\$10,345-\$87,309) | \$62,834<br>(\$81,172) |
| *Without ICU or IMV*<br>*N=2208* | 51<br>(2.3) | 3<br>(1-4) | 5.0<br>(7.9) | \$6964<br>(\$3486-\$10,644) | \$13,555<br>(\$24,868) | \$26,824<br>(\$9351-\$71,001) | \$54,737<br>(\$75,877) |
| *With ICU, but without IMV*<br>*N=518* | 14<br>(2.7) | 4<br>(2-8) | 5.5<br>(4.7) | \$12,263<br>(\$5375-\$23,383) | \$20,603<br>(\$25,093) | \$41,664<br>(\$12,241-\$93,515) | \$55,615<br>(\$53,318) |
| *Without ICU, but with IMV*<br>*N=18* | 0<br>(0) | - | - | - | - | - | - |
| *With ICU and IMV*<br>*N=190* | 5<br>(2.6) | 4<br>(3-23) | 11.6<br>(12.4) | \$20,911<br>(\$10,983-\$75,091) | \$42,482<br>(\$40,564) | \$123,614<br>(\$80,954-\$242,950) | \$165,632<br>(\$134,993) |
|  | Index hospitalizations:<br>May 2021-Jun 2021 |  |  |  |  |  |  |
|  | N | Hospital LOS, days |  | Hospital costs<sup>a</sup> |  | Hospital charges |  |
| Index hospitalization status | (%) | Median (Q1-Q3) | Mean (SD) | Median (Q1-Q3) | Mean (SD) | Median (Q1-Q3) | Mean (SD) |
| **All inpatient admissions**<br>**N=233** | 4<br>(1.7) | 2<br>(1.5-5) | 3.3<br>(3.2) | \$10,756<br>(\$5156-\$24,081) | \$13,331<br>(\$9722) | \$17,305<br>(\$10,792-\$79,828) | \$45,310<br>(63,020) |
| *Without ICU or IMV*<br>*N=177* | 4<br>(2.3) | 2<br>(1.5-5) | 3.3<br>(3.2) | \$10,756<br>(\$5156-\$24,081) | \$13,331<br>(\$9722) | \$17,305<br>(\$10,792-\$79,828) | \$45,310<br>(63,020) |
| *With ICU, but without IMV*<br>*N=38* | 0<br>(0) | - | - | - | - | - | - |
| *Without ICU, but with IMV*<br>*N=1* | 0<br>(0) | - | - | - | - | - | - |
| *With ICU and IMV* | 0 | - | - | - | - | - | - |

| *N=17* | (0) |  |  |  |  |  |  |
| --- | --- | --- | --- | --- | --- | --- | --- |
|  | Index<br>hospitalizations:<br>Jul 2021-Sep 2021 | Hospital LOS, days |  | Hospital costs<sup>a</sup> |  | Hospital charges |  |
|  | N<br>(%)<sup>b</sup> | Median<br>(Q1-Q3) | Mean<br>(SD) | Median<br>(Q1-Q3) | Mean<br>(SD) | Median<br>(Q1-Q3) | Mean<br>(SD) |
| **Index hospitalization status** |  |  |  |  |  |  |  |
| **All inpatient admissions** | 22 | 3 | 3.6 | \$4544 | \$9274 | \$34,485 | \$38,656 |
| **N=1406** | (1.6) | (2-5) | (2.5) | (\$3204-\$10,765) | (\$9229) | (\$17,703-\$52,692) | (\$27,520) |
| ***Without ICU or IMV*** | 19 | 3 | 3.7 | \$4544 | \$9256 | \$37,594 | \$38,694 |
| **N=1096** | (1.7) | (2-5) | (2.6) | (\$3034-\$10,765) | (\$9584) | (\$19,696-\$52,692) | (\$25,214) |
| ***With ICU, but without IMV*** | 2 | 2 | 2 | \$3204 | \$3204 | \$11,168 | \$11,168 |
| **N=201** | (1.0) | (2-2) | (0) | - | - | (\$9533-\$12,803) | (\$2312) |
| ***Without ICU, but with IMV*** | 1 | 4 | 4 | \$15,614 | \$15,614 | \$92,917 | \$92,917 |
| **N=15** | (6.7) | - | - | - | - | - | - |
| ***With ICU and IMV*** | 0 |  |  |  |  |  |  |
| **N=94** | (0) | - | - | - | - | - | - |

<sup>a</sup> Among patients with validated costs >\$0.
<sup>b</sup> Percentage among those with COVID-19-associated index hospitalizations, overall and stratified by the 4 COVID-19 disease progression states.
ICU: Intensive care unit; IMV: Invasive mechanical ventilation; LOS: Length of stay; PHD SR: Premier Healthcare Database Special Release; Q1: First quartile; Q3: Third quartile; SD: Standard deviation

**Figure 2.**
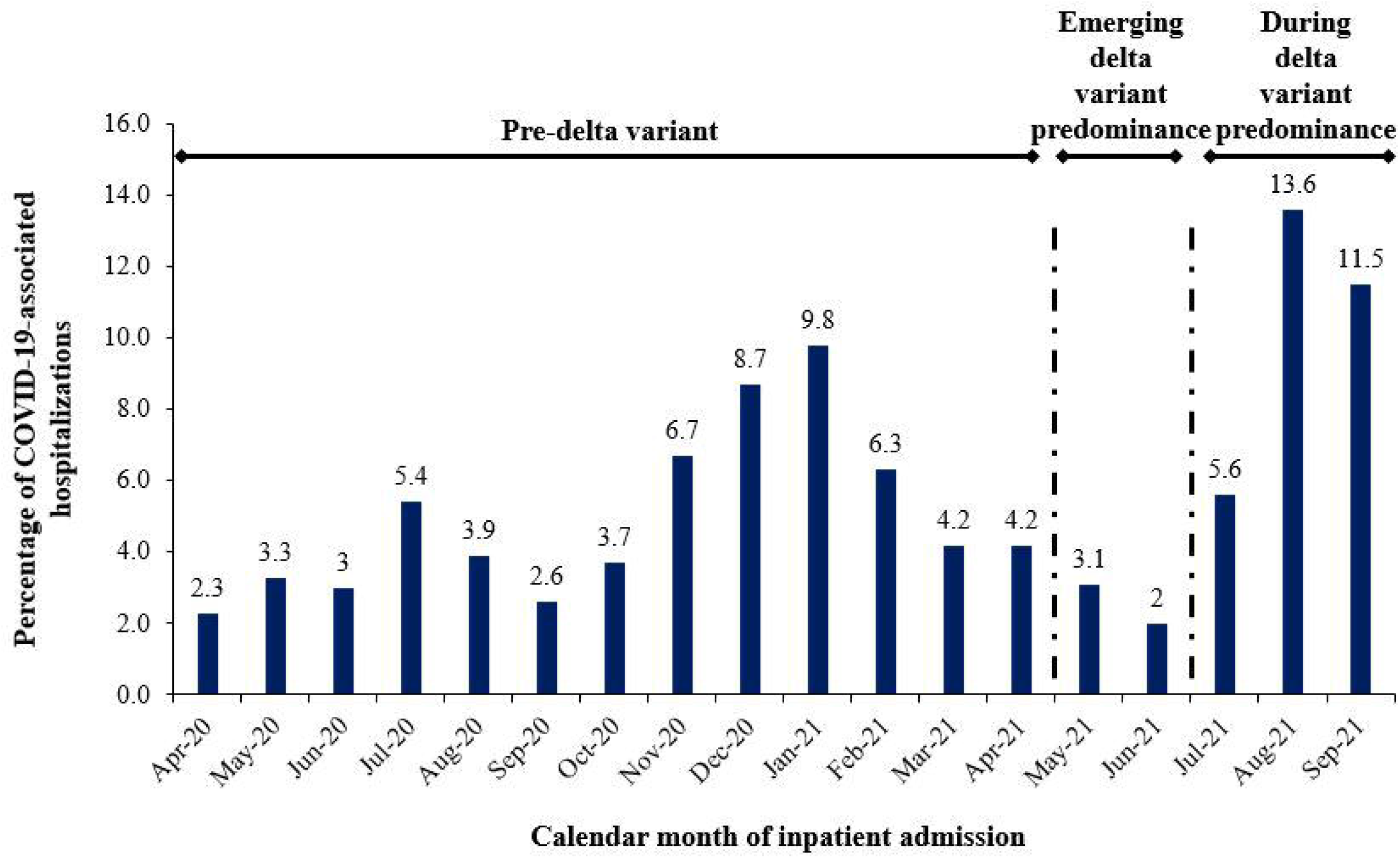
PHD SR: Frequency of index hospitalizations by calendar month among children aged 0-11 years

### Sensitivity analysis: HV RTIE database

Of the total 3,263,406 inpatient admissions reported in HV RTIE database between 1 April 2020 and 30 September 2021, 10.0% were children aged ≤11 years. Of those, 0.6% (N=1,962) had a COVID-19-associated hospitalization with a recorded primary or secondary diagnosis. Among these 1,962 pediatric patients, 63.5% (1,246) were aged 0-4 years and the remaining 36.5% (716) were aged 5-11 years. Supplementary Tables 3-10 present the demographics, comorbidities, health outcomes and associated costs, and readmission outcomes of the study population selected from the HV RTIE database.

Among the study population aged 0-11 years, a majority (75.5%) were admitted to the general ward, 432 (22.0%) were admitted to an ICU, and 136 (6.9%) received IMV. As shown in Supplementary Figure 1, the frequency of COVID-19-associated hospitalization declined from January 2021 to June 2021, but then increased again in July 2021 during delta predominance, which was consistent with the trend observed among the pediatric population selected from the PHD SR.

The findings of the sensitivity analysis using the HV RTIE database were generally consistent with the main analysis conducted using the PHD SR. Major similarities included the distribution of the 0-4 and 5-11 year age groups with COVID-19-associated hospitalizations, the prevalence of comorbid conditions, and the increasing median hospital LOS and costs with greater COVID-19 severity. Additionally, based on ICU admission, COVID-19 severity was greater among those aged 5-11 years, with 26.5% admitted to the ICU compared to 19.4% among children aged 0-4 years. The COVID-19 readmission rate was similar (2.6% versus 2.1%), as well as the proportion of all-cause readmissions that were COVID-19-associated readmissions (29.7% versus 32.3%).

The most notable differences included that the proportion of pediatric COVID-19 cases admitted to rural hospitals was higher in HV RTIE database than in the PHD SR (12.0% versus 6.1%), the proportion in the no ICU but IMV cohort was higher (2.5% versus 0.7%), but the proportion in the ICU and IMV cohort was lower (4.4% versus 6.6%). The median hospital LOS (2 versus 3 days) was longer, while median hospital costs ($10,286 versus $6,164) were approximately 67% higher compared to the findings using the PHD SR.

### Sensitivity analysis: Patients with a primary diagnosis of COVID-19

Of the 4,573 patients aged 0-11 years with a COVID-19-associated index hospitalization, 56.0% (N=2,561) had a COVID-19 primary diagnosis. The distribution of patients across the disease progression states was very similar to the main analyses; while the measured outcomes were slightly lower, consistent with a prior study [7] (data available upon request).

## Discussion

The COVID-19 pandemic has represented an unprecedented challenge for hospitals and healthcare systems. Using the large PHD SR as a data source, we assessed hospitalization patterns in a pediatric population. We identified 4,573 patients aged 0-11 years with COVID-19-associated hospitalizations during April 2020 through September 2021. Of those, almost one-quarter (23.1%) were admitted to the ICU and 7.3% had IMV usage. A majority (68.0%) of the COVID-19-associated hospitalizations were among children 0-4 years of age (mean age: 0.8 years). However, based on ICU admission, COVID-19 severity was greater among those aged 5-11 years (mean age: 8.3 years), with 30.3% admitted to the ICU compared to 19.8% among children aged 0-4 years. Similarly, other severity outcomes were less favorable among children 5-11 years, including longer hospital LOS, higher hospital costs as well as higher readmission rates. These findings were relatively unchanged when limited to hospitalized pediatric COVID-19 cases with a primary diagnosis and when examined using the HV RTIE database as a data source.

The higher frequency of COVID-19-associated hospitalizations in the youngest age group (0-4 years) versus those 5-11 years of age is consistent with COVID-NET surveillance data [1,2]. In the COVID-NET dataset, cumulative rates of COVID-19-associated hospitalizations were nearly two times higher among children aged 0-4 years than among those aged 5-11 years [1], whereas, in the study of Kim et al. [6], 71% of the COVID-19-associated hospitalizations among those 0-11 years of age were of children aged 0-4 years.

The observed hospital resource use and in-hospital mortality rates among the pediatric study population herein were also relatively similar to those reported in the three CDC studies of pediatric COVID-19-associated hospitalizations [2,3,6]. In the earlier (1 March 2020 through 25 July 2020) study of Kim et al. [6], which utilized COVID-NET surveillance data, of 208 children and adolescents (0-17 years of age) with complete medical chart reviews, 33.2% had an ICU admission, 5.8% had IMV usage, and 0.5% (N=1) died; median hospital LOS was 2.5 days [6]. In the study by Delahoy et al. [2], which also utilized COVID-NET surveillance data, of the 3,116 children and adolescents, aged 0-17 years, hospitalized with COVID-19 during 1 March 2020 through 19 June 2021 (with complete clinical data available), 26.5% were admitted to an ICU, 6.1% required IMV, and 0.7% (N=21) died; the median hospital LOS was 3 days. In the study of Siegel et al. [3], which utilized the BD Insights Research Database, among the pediatric study population with COVID-19-associated hospitalizations during August 2020 through June 2021 (aged 0-17 years; N=1790), ICU admissions rates ranged from 10% to 25% and IMV usage ranged from 0% to 3%; median hospital LOS ranged 2 to 3 days. The frequency of IMV usage among the pediatric patients with COVID-19-associated hospitalization in the study of Siegel et al. [3] was lower than observed in our study and also that observed by Delahoy et al. [2]; this may be related to the contributing hospitals represented in the BD Insights Research Database, which includes three children’s hospitals with the remainder being community hospitals [3].

In both the studies of Delahoy et al. [2] and Siegel et al. [3], pediatric COVID-19-associated hospitalization rates increased during the period of emerging delta variant predominance, up to 5-fold among children and adolescents aged 0-17 years and up to 10-fold among those 0-4 years of age, which is similar to the findings of this study. Also similar to our observations regarding ICU admission and IMV usage rates, Delahoy et al. [2] did not find a significant increase in ICU admission during delta variant predominance (26.5%: 20 June –31 July 2021) compared to the earlier period (23.2%: 1 March 2020–19 June 2021) and while in this study, the proportion with IMV usage increased from 6.1% to 9.8%, it was based on a small number of observations and was also not found to be statistically significant. However, Siegel et al. [3] did find that ICU admission and IMV usage increased in July and August 2021 versus June 2021 among children and adolescents, 0-17 years of age [3]. Whether the increases in hospitalization rates among pediatric populations during July, August, and September of 2021 in the US were related to the predominance of the delta variant and its potentially increased transmission and virulence or other variables (e.g., increased transmission due to relaxing of masking requirements, in-person school re-opening, greater social interaction, etc.) could not be discerned from the two CDC surveillance studies [2,3] or our current study, and further follow-up studies are needed.

Di Fusco et al. [7] reported on 1,671 pediatric patients (0-4 years: 35.7%; 5-17: 64.3%) with COVID-19-associated hospitalizations during April 1 through October 31, 2020. A similar median hospital LOS of 2 days among children aged 0-4 years was observed; however, median hospital costs were $6,012 compared to $5,157 in the current study, suggestive of a possible decline in COVID-19-associated hospitalization costs across all such inpatient admissions. Additionally, the median hospitals costs of pediatric patients aged 0-11 years in our study, were approximately 50% lower than found for adults ($12,046), which is partly due to the shorter hospital LOS of pediatric patients compared to adults hospitalized for COVID-19 (2 vs 5 days, respectively) [7].

To our knowledge, this is the first study to evaluate longer-term hospitalization outcomes in a pediatric population hospitalized with COVID-19 in the US. In the main analyses and HV RTIE database sensitivity analyses, respectively, 2.1% and 2.6% had a COVID-19-associated readmission. These readmissions represented up to one-third of all-cause readmissions and stress the long-term and severe complications experienced by some pediatric patients hospitalized for COVID-19.

From our dataset of pediatric COVID-19-associated hospitalizations over the three study periods, median hospital costs appear to have trended downward from April 2020-April 2021 to July-September 2021 (median: $6,357 versus $5,433, respectively) but hospital costs were highest during May and June 2021 (median: $7,449). Hospital costs among pediatric patients admitted to the ICU may have also decreased in the most recent time period of this study, but IMV usage remains costly. Such trends in declining costs for pediatric COVID-19-associated hospitalizations are consistent with that of adult COVID-19-associated hospitalizations. Ohsfeldt et al. [17], found that among 247,590 adult COVID-19-associated hospitalizations during April through December of 2020, median hospital costs declined by 50% and median ICU costs declined by 35%. They proposed that the declining trend in hospital costs may be related to decreased hospital resource use, which itself may be related to improved understanding and treatment of patients hospitalized with COVID-19 [17]. Because the frequency of ICU admission and IMV usage remained relatively stable over the entire study period among the pediatric population in this study, this may have led to a lesser decline in COVID-19-associated pediatric hospital costs compared to adults.

The findings of this study underscore the clinical and economic value of comprehensive vaccination of pediatric populations, especially in the context of the limited treatment options recommended for children hospitalized with COVID-19 [18] and emergence of more transmissible and/or virulent SARS-CoV-2 variants. A retrospective single-center pediatric cohort study in Chicago, IL found that between October 15, 2020 and August 31, 2021, among children and adolescents (≤18 years of age), the gamma variant was associated with greater COVID-19 illness severity, as determined by increased rates of hospitalization, respiratory support, and World Health Organization Clinical Progression Scale score ≥6, while the increased pediatric hospitalization rate observed during the delta variant predominance was associated with increased transmission [19]. Delahoy et al. [2] reported that COVID-19 vaccination reduced the risk of COVID-19-associated hospitalization by approximately 10-fold among adolescents aged 12-17 years, relative to the unvaccinated in this age group, which was the only age group eligible for vaccination during the time period of the study. As the COVID-19 pandemic continues throughout the world with potential seasonal resurgences of transmission and the emergence of more transmissible and/or virulent SARS-CoV-2 variants, such as Omicron, it is critical to examine the risks and benefits of vaccination for all age groups, as well as the potential longer term clinical and economic benefits of COVID-19 vaccination.

As of 2 December 2021, the number of pediatric cases of COVID-19 illness totaled 7,032,612 in the US [20]; and as of 5 December 2021, only approximately 17% of children, 5-11 years of age, had received at least one COVID-19 vaccine dose, representing nearly 4.8 million children in this age group of 28 million [21]. Thus, the start of vaccine rollout in this age group has been rather slow, potentially impacted by parental hesitancy [21]. With these perspectives, the findings of this study emphasize the importance of COVID-19 vaccination of the pediatric population, which, in the US, represents 22.2% of the overall population [20].

## Limitations

The findings of this study should be interpreted in the context of the following limitations. Only in-hospital resource use and costs were assessed, and additional analyses that include care in outpatient settings, as well as indirect costs (e.g., lost employment income, transit or transport costs, childcare costs, etc.) are needed to provide a more complete description of the clinical and economic burdens of pediatric patients hospitalized with COVID-19. Moreover, while readmission outcomes allowed early insights into long-term complications, the long-term sequalae of COVID-19 illness, which have been reported in some children [4,5], were not evaluated in this study.

COVID-19 illness in hospitalized pediatric patients was identified using the diagnosis code U07.1 only and was not laboratory-confirmed. However, Kadri et al. [13] analyzed 51,907 inpatient discharges with confirmatory laboratory testing data and reported that the diagnosis code U07.1, had a 91.5% positive predictive value (9.6% positive SARS-CoV-2-PCR test results; sensitivity: 98.0%; specificity: 99.0%) for identifying COVID-19 cases (1 April-31 May 2020). In this study, we also required a COVID-19 diagnosis to be POA. This more stringent requirement may have led to an underestimation of hospitalized COVID-19 pediatric cases since children who may have been admitted to the hospital for another cause may have been incidentally found to be COVID-19 positive. However, some of such cases may have been captured with the allowed secondary diagnosis of COVID-19 in the main analyses. Regarding the reported hospital costs, among the overall pediatric study population, 0-11 years of age, approximately 17% (N=791) did not have validated cost information recorded in the PHD SR. Therefore, the reported hospital costs are reflective of only the subset of hospitalized pediatric patients with such hospital cost information. In the PHD SR, COVID-19-associated hospital readmissions may be underestimated since this information is only captured when patients were readmitted to the same hospital (or hospital system) within the database.

The PHD SR database contains limited patient medical history, particularly among pediatric patients; thus, the prevalence of comorbid conditions among this pediatric study population may have been underestimated. While the PHD SR database contains information on approximately one-quarter of hospital admissions across the US, it is possible that our study findings may not be representative of the entire population of hospitalized COVID-19 pediatric patients in the US, nor those populations in areas and geographic regions with less representation of contributing hospitals in the data source (e.g., rural [27%] versus urban [73%]; South [44%] versus West [14.5%]). Additionally, it is possible that the hospitals contributing to the PHD SR and the HV RTIE database may overlap; however, de-duplicating patients is not possible due to the de-identified nature of the databases. We chose to use the HV RTIE database as a sensitivity analysis to verify trends observed among the pediatric population selected from the PHD SR, since the HV RTIE database contained fewer hospitalized pediatric patients. Lastly, billing and coding errors could have occurred.

## Conclusions

The findings from this retrospective analysis using a large US hospital-based, all-payer database underscore that children aged 0-11 years can experience severe COVID-19 illness requiring hospitalization and substantial hospital resource use, further heightening the clinical and healthcare economic burdens of the continuing COVID-19 pandemic. This study, to our knowledge, is the first to provide detailed health and economic outcomes for the pediatric population, 0-11 years of age, in the US, and additionally to evaluate COVID-19-associated readmissions patterns among this age group. Our conclusions support recent US Food & Drug Administration authorization and CDC recommendations to offer COVID-19 vaccination to children aged 5-11 years and highlight the need for vigilant efforts to prevent COVID-19 illness in the overall pediatric population.

## Data Availability

Data generated or analyzed during this study are available upon request.

## Transparency

### Declaration of financial/other relationships

MDF, MMM, JEA, DM, TSS, JLN, AC, and LJM are employees of Pfizer. JL is an employee of Novosys Health, which received financial support from Pfizer in connection with the study and the development of this manuscript. SV is an external consultant for Pfizer who has received consulting fees from Pfizer in connection with the study and the development of this manuscript.

### Author contributions

All named authors meet the International Committee of Medical Journal Editors (ICMJE) criteria for authorship for this article. All authors contributed to study conception and design, data acquisition, analysis, and interpretation, drafting and revising of the manuscript, and have given their approval for this manuscript version to be published.

## Acknowledgements

The authors acknowledge Tamuno Alfred and Maya Reimbaeva (Pfizer employees) and Andy Surinach (Genesis Research employee) for specific contributions to this research project. Editorial support was provided by Melissa Lingohr-Smith at Novosys Health and was funded by Pfizer.

## Data availability statement

Data generated or analyzed during this study are available upon request.

