## supplementary for "COVID-19-Associated Hospitalizations Among Children Less Than 12 Years of Age in the United States"

**Supplementary material**

**Supplementary Table 1. PHD SR: Demographics, hospitalization characteristics, and comorbid conditions among children aged 0-4 years**

|  | **Overall study population  N=3111 (100%)** | | **Without ICU**  **or IMV  N=2472 (79.5%)** | | **With ICU, but without IMV  N=410 (13.2%)** | | **Without ICU, but with IMV  N=24 (0.8%)** | | **With ICU and IMV N=205 (6.6%)** | |
| --- | --- | --- | --- | --- | --- | --- | --- | --- | --- | --- |
| ***Age (years)*** |  |  |  |  |  |  |  |  |  |  |
| Median, Q1-Q3 | 0 | 0-1 | 0 | 0-1 | 1 | 0-2 | 1 | 0-3 | 0 | 0-2 |
| Mean, SD | 0.8 | 1.3 | 0.8 | 1.2 | 1.2 | 1.3 | 1.4 | 1.6 | 1.1 | 1.4 |
|  | ***N*** | ***%*** | ***N*** | ***%*** | ***N*** | ***%*** | ***N*** | ***%*** | ***N*** | ***%*** |
| ***Sex*** |  |  |  |  |  |  |  |  |  |  |
| Male | 1756 | 56.4 | 1,367 | 55.3 | 264 | 64.4 | 16 | 66.7 | 109 | 53.2 |
| Female | 1355 | 43.6 | 1,105 | 44.7 | 146 | 35.6 | 8 | 33.3 | 96 | 46.8 |
| ***Race/ethnicity*** |  |  |  |  |  |  |  |  |  |  |
| White non-Hispanic | 807 | 25.9 | 659 | 26.7 | 88 | 21.5 | 11 | 45.8 | 49 | 23.9 |
| Black non-Hispanic | 471 | 15.1 | 365 | 14.8 | 68 | 16.6 | 1 | 4.2 | 37 | 18.0 |
| Asian non-Hispanic | 64 | 2.1 | 56 | 2.3 | 6 | 1.5 | 0 | 0.0 | 2 | 1.0 |
| Other non-Hispanic | 186 | 6.0 | 140 | 5.7 | 21 | 5.1 | 5 | 20.8 | 20 | 9.8 |
| White Hispanic | 562 | 18.1 | 440 | 17.8 | 85 | 20.7 | 3 | 12.5 | 34 | 16.6 |
| Black Hispanic | 28 | 0.9 | 24 | 1.0 | 3 | 0.7 | 0 | 0.0 | 1 | 0.5 |
| Asian Hispanic | 2 | 0.1 | 2 | 0.1 | 0 | 0.0 | 0 | 0.0 | 0 | 0.0 |
| Other Hispanic | 348 | 11.2 | 278 | 11.2 | 44 | 10.7 | 2 | 8.3 | 24 | 11.7 |
| Unknown^a^ | 643 | 20.7 | 508 | 20.6 | 95 | 23.2 | 2 | 8.3 | 38 | 18.5 |
| ***Insurance type*** |  |  |  |  |  |  |  |  |  |  |
| Commercial | 689 | 22.1 | 572 | 23.1 | 76 | 18.5 | 4 | 16.7 | 37 | 18.0 |
| Medicaid | 2119 | 68.1 | 1668 | 67.5 | 287 | 70.0 | 18 | 75.0 | 146 | 71.2 |
| Medicare | 8 | 0.3 | 6 | 0.2 | 1 | 0.2 | 0 | 0.0 | 1 | 0.5 |
| Other | 220 | 7.1 | 172 | 7.0 | 36 | 8.8 | 1 | 4.2 | 11 | 5.4 |
| Uninsured | 75 | 2.4 | 54 | 2.2 | 10 | 2.4 | 1 | 4.2 | 10 | 4.9 |
| ***Hospital Census region*** |  |  |  |  |  |  |  |  |  |  |
| Northeast | 434 | 14.0 | 364 | 14.7 | 40 | 9.8 | 2 | 8.3 | 28 | 13.7 |
| Midwest | 426 | 13.7 | 343 | 13.9 | 54 | 13.2 | 2 | 8.3 | 27 | 13.2 |
| South | 1693 | 54.4 | 1316 | 53.2 | 253 | 61.7 | 14 | 58.3 | 110 | 53.7 |
| West | 558 | 17.9 | 449 | 18.2 | 63 | 15.4 | 6 | 25.0 | 40 | 19.5 |
| ***Hospital population served*** |  |  |  |  |  |  |  |  |  |  |
| Urban | 2918 | 93.8 | 2,338 | 94.6 | 365 | 89.0 | 18 | 75.0 | 197 | 96.1 |
| Rural | 193 | 6.2 | 134 | 5.4 | 45 | 11.0 | 6 | 25.0 | 8 | 3.9 |
| ***COVID-19 diagnosis position*** |  |  |  |  |  |  |  |  |  |  |
| Primary | 1829 | 58.8 | 1,509 | 61.0 | 227 | 55.4 | 12 | 50.0 | 81 | 39.5 |
| Secondary | 1282 | 41.2 | 963 | 39.0 | 183 | 44.6 | 12 | 50.0 | 124 | 60.5 |
| ***Calendar period of admission*** |  |  |  |  |  |  |  |  |  |  |
| April 2020-April 2021 | 1969 | 63.3 | 1,562 | 63.2 | 276 | 67.3 | 11 | 45.8 | 120 | 58.5 |
| May 2021-June 2021 | 143 | 4.6 | 110 | 4.4 | 19 | 4.6 | 1 | 4.2 | 13 | 6.3 |
| July 2021-September 2021 | 999 | 32.1 | 800 | 32.4 | 115 | 28.0 | 12 | 50.0 | 72 | 35.1 |
| ***Discharge status^b^*** |  |  |  |  |  |  |  |  |  |  |
| Home/self-care | 2951 | 94.9 | 2394 | 96.8 | 390 | 95.1 | 15 | 62.5 | 152 | 74.1 |
| Expired/died | 14 | 0.5 | 0 | 0.0 | 0 | 0.0 | 0 | 0.0 | 14 | 6.8 |
| Transferred to other facility | 81 | 2.6 | 48 | 1.9 | 11 | 2.7 | 6 | 25.0 | 16 | 7.8 |
| ***Comorbid conditions*** |  |  |  |  |  |  |  |  |  |  |
| Number of comorbidities: 0 | 2153 | 69.2 | 1811 | 73.2 | 233 | 56.8 | 16 | 66.7 | 93 | 45.4 |
| Number of comorbidities: 1-2 | 896 | 28.8 | 627 | 25.4 | 167 | 40.7 | 8 | 33.3 | 94 | 45.9 |
| Number of comorbidities: ≥3 | 62 | 1.9 | 34 | 1.4 | 10 | 2.4 | 0 | 0.0 | 18 | 8.8 |
| Median, Q1-Q3 | 0 | 0-1 | 0 | 0-1 | 0 | 0-1 | 0 | 0-1 | 1 | 0-1 |
| Mean, SD | 0.4 | 0.7 | 0.4 | 0.7 | 0.6 | 0.8 | 0.4 | 0.6 | 0.9 | 1.1 |
| ***Comorbid conditions*** |  |  |  |  |  |  |  |  |  |  |
| Diabetes | 21 | 0.7 | 10 | 0.4 | 10 | 2.4 | 0 | 0.0 | 1 | 0.5 |
| Obesity/overweight | 22 | 0.7 | 15 | 0.6 | 2 | 0.5 | 0 | 0.0 | 5 | 2.4 |
| Hypertension | 58 | 1.9 | 23 | 0.9 | 18 | 4.4 | 0 | 0.0 | 17 | 8.3 |
| CKD or ESRD | 95 | 3.1 | 49 | 2.0 | 21 | 5.1 | 0 | 0.0 | 25 | 12.2 |
| Neurological disease | 167 | 5.4 | 86 | 3.5 | 26 | 6.3 | 4 | 16.7 | 51 | 24.9 |
| Psychiatric disorders | 7 | 0.2 | 1 | 0.0 | 3 | 0.7 | 0 | 0.0 | 3 | 1.5 |
| Malignancy (solid or hematologic) | 98 | 3.2 | 79 | 3.2 | 14 | 3.4 | 0 | 0.0 | 5 | 2.4 |
| Asthma/reactive airway disease | 180 | 5.8 | 127 | 5.1 | 37 | 9.0 | 0 | 0.0 | 16 | 7.8 |
| Chronic liver disease | 1 | 0.0 | 0 | 0.0 | 1 | 0.2 | 0 | 0.0 | 0 | 0.0 |
| Congenital heart condition | 23 | 0.7 | 11 | 0.4 | 6 | 1.5 | 0 | 0.0 | 6 | 2.9 |
| Congenital lung condition | 5 | 0.2 | 2 | 0.1 | 2 | 0.5 | 0 | 0.0 | 1 | 0.5 |
| Down syndrome/chromosomal anomaly | 41 | 1.3 | 23 | 0.9 | 10 | 2.4 | 1 | 4.2 | 7 | 3.4 |
| Immunocompromised^c^ | 605 | 19.4 | 444 | 18.0 | 103 | 25.1 | 3 | 12.5 | 55 | 26.8 |
| Autoimmune disease | 18 | 0.6 | 8 | 0.3 | 10 | 2.4 | 0 | 0.0 | 0 | 0.0 |
| Sickle cell | 59 | 1.9 | 50 | 2.0 | 6 | 1.5 | 0 | 0.0 | 3 | 1.5 |
| Disability^d^ | 19 | 0.6 | 10 | 0.4 | 2 | 0.5 | 1 | 4.2 | 6 | 2.9 |
| Transplant (bone marrow and organ) | 19 | 0.6 | 15 | 0.6 | 1 | 0.2 | 1 | 4.2 | 2 | 1.0 |

^a^ Unknown refers to either one of, or both, race and ethnicity are unknown.

^b^ Reported only top three discharge status here, remaining such as skilled nursing facility, home health organization, hospice, rehabilitation facility, and/or other are not listed.

^c^ Immunocompromised conditions included HIV/AIDS, malignancy, transplants, rheumatologic/other inflammatory conditions, primary immunodeficiency, CKD/ESRD, and other immune conditions.

^d^ Includes neurologic, neurodevelopmental, intellectual, physical, vision or hearing impairment.

AIDS: Acquired immunodeficiecy syndrome. CKD: Chronic kidney disease; ESRD: End stage renal disease; HIV: Human immunodeficiency virus; ICU: Intensive care unit; IMV: Invasive mechanical ventilation; PHD SR: Premier Healthcare Database Special Release; Q1: First quartile; Q3: Third quartile; SD: Standard deviation.

**Supplementary Table 2. PHD SR: Demographics, hospitalization characteristics, and comorbid conditions among children aged 5-11 years**

|  | **Overall study population  N=1462 (100%)** | | **Without ICU**  **or IMV  N=1009 (69.0%)** | | **With ICU, but without IMV  N=347 (23.7%)** | | **Without ICU, but with IMV  N=10 (0.7%)** | | **With ICU and IMV N=96 (6.6%)** | |
| --- | --- | --- | --- | --- | --- | --- | --- | --- | --- | --- |
| ***Age (years)*** |  |  |  |  |  |  |  |  |  |  |
| Median, Q1-Q3 | 8 | 7-10 | 8 | 7-10 | 8 | 7-10 | 9 | 8-10 | 9 | 7-10 |
| Mean, SD | 8.3 | 2.1 | 8.3 | 2.1 | 8.3 | 2.0 | 8.7 | 1.6 | 8.4 | 2.0 |
|  | ***N*** | ***%*** | ***N*** | ***%*** | ***N*** | ***%*** | ***N*** | ***%*** | ***N*** | ***%*** |
| ***Sex*** |  |  |  |  |  |  |  |  |  |  |
| Male | 804 | 55.0 | 554 | 54.9 | 191 | 55.0 | 3 | 30.0 | 56 | 58.3 |
| Female | 658 | 45.0 | 455 | 45.1 | 156 | 45.0 | 7 | 70.0 | 40 | 41.7 |
| ***Race/ethnicity*** |  |  |  |  |  |  |  |  |  |  |
| White non-Hispanic | 387 | 26.5 | 290 | 28.7 | 72 | 20.7 | 4 | 40.0 | 21 | 21.9 |
| Black non-Hispanic | 287 | 19.6 | 184 | 18.2 | 78 | 22.5 | 2 | 20.0 | 23 | 24.0 |
| Asian non-Hispanic | 42 | 2.9 | 29 | 2.9 | 6 | 1.7 | 1 | 10.0 | 6 | 6.3 |
| Other non-Hispanic | 70 | 4.8 | 48 | 4.8 | 18 | 5.2 | 0 | 0.0 | 4 | 4.2 |
| White Hispanic | 233 | 15.9 | 155 | 15.4 | 62 | 17.9 | 1 | 10.0 | 15 | 15.6 |
| Black Hispanic | 20 | 1.4 | 14 | 1.4 | 6 | 1.7 | 0 | 0.0 | 0 | 0.0 |
| Other Hispanic | 193 | 13.2 | 139 | 13.8 | 42 | 12.1 | 1 | 10.0 | 11 | 11.5 |
| Unknown^a^ | 230 | 15.7 | 150 | 14.9 | 63 | 18.2 | 1 | 10.0 | 16 | 16.7 |
| ***Insurance type*** |  |  |  |  |  |  |  |  |  |  |
| Commercial | 397 | 27.2 | 288 | 28.5 | 87 | 25.1 | 2 | 20.0 | 20 | 20.8 |
| Medicaid | 953 | 65.2 | 643 | 63.7 | 234 | 67.4 | 8 | 80.0 | 68 | 70.8 |
| Medicare | 7 | 0.5 | 6 | 0.6 | 1 | 0.3 | 0 | 0.0 | 0 | 0.0 |
| Other | 81 | 5.5 | 58 | 5.7 | 15 | 4.3 | 0 | 0.0 | 8 | 8.3 |
| Uninsured | 24 | 1.6 | 14 | 1.4 | 10 | 2.9 | 0 | 0.0 | 0 | 0.0 |
| ***Hospital Census region*** |  |  |  |  |  |  |  |  |  |  |
| Northeast | 234 | 16.0 | 162 | 16.1 | 49 | 14.1 | 2 | 20.0 | 21 | 21.9 |
| Midwest | 172 | 11.8 | 121 | 12.0 | 37 | 10.7 | 4 | 40.0 | 10 | 10.4 |
| South | 818 | 56.0 | 549 | 54.4 | 215 | 62.0 | 3 | 30.0 | 51 | 53.1 |
| West | 238 | 16.3 | 177 | 17.5 | 46 | 13.3 | 1 | 10.0 | 14 | 14.6 |
| ***Hospital population served*** |  |  |  |  |  |  |  |  |  |  |
| Urban | 1376 | 94.1 | 966 | 95.7 | 310 | 89.3 | 10 | 100.0 | 90 | 93.8 |
| Rural | 86 | 5.9 | 43 | 4.3 | 37 | 10.7 | 0 | 0.0 | 6 | 6.3 |
| ***COVID-19 diagnosis position*** |  |  |  |  |  |  |  |  |  |  |
| Primary | 732 | 50.1 | 485 | 48.1 | 189 | 54.5 | 5 | 50.0 | 53 | 55.2 |
| Secondary | 730 | 49.9 | 524 | 51.9 | 158 | 45.5 | 5 | 50.0 | 43 | 44.8 |
| ***Calendar period of admission*** |  |  |  |  |  |  |  |  |  |  |
| April 2020-April 2021 | 965 | 66.0 | 646 | 64.0 | 242 | 69.7 | 7 | 70.0 | 70 | 72.9 |
| May 2021-June 2021 | 90 | 6.2 | 67 | 6.6 | 19 | 5.5 | 0 | 0.0 | 4 | 4.2 |
| July 2021-September 2021 | 407 | 27.8 | 296 | 29.3 | 86 | 24.8 | 3 | 30.0 | 22 | 22.9 |
| ***Discharge status^b^*** |  |  |  |  |  |  |  |  |  |  |
| Home/self-care | 1347 | 92.1 | 945 | 93.7 | 329 | 94.8 | 7 | 70.0 | 66 | 68.8 |
| Expired/died | 9 | 0.6 | 0 | 0.0 | 1 | 0.3 | 1 | 10.0 | 7 | 7.3 |
| Transferred to other facility | 63 | 4.3 | 49 | 4.9 | 9 | 2.6 | 1 | 10.0 | 4 | 4.2 |
| ***Comorbid conditions*** |  |  |  |  |  |  |  |  |  |  |
| Number of comorbidities: 0 | 410 | 28.0 | 355 | 35.2 | 44 | 12.7 | 2 | 20 | 9 | 9.4 |
| Number of comorbidities: 1-2 | 896 | 61.3 | 583 | 57.8 | 255 | 73.5 | 5 | 50 | 53 | 55.2 |
| Number of comorbidities: ≥3 | 156 | 10.7 | 71 | 7.0 | 48 | 13.8 | 3 | 30 | 34 | 35.4 |
| Median, Q1-Q3 | 1 | 0-2 | 1 | 0-2 | 1 | 1-2 | 2 | 1-3 | 2 | 1-3 |
| Mean, SD | 1.2 | 1.0 | 1.0 | 1.0 | 1.5 | 0.9 | 1.7 | 1.2 | 2.1 | 1.3 |
| ***Comorbid conditions*** |  |  |  |  |  |  |  |  |  |  |
| Diabetes | 62 | 4.2 | 20 | 2.0 | 40 | 11.5 | 0 | 0.0 | 2 | 2.1 |
| Obesity/overweight | 168 | 11.5 | 95 | 9.4 | 51 | 14.7 | 3 | 30.0 | 19 | 19.8 |
| Hypertension | 50 | 3.4 | 26 | 2.6 | 10 | 2.9 | 0 | 0.0 | 14 | 14.6 |
| CKD or ESRD | 146 | 10.0 | 51 | 5.1 | 57 | 16.4 | 1 | 10.0 | 37 | 38.5 |
| Neurological disease | 184 | 12.6 | 113 | 11.2 | 43 | 12.4 | 4 | 40.0 | 24 | 25.0 |
| Psychiatric disorders | 92 | 6.3 | 70 | 6.9 | 14 | 4.0 | 1 | 10.0 | 7 | 7.3 |
| Malignancy (solid or hematologic) | 80 | 5.5 | 63 | 6.2 | 14 | 4.0 | 0 | 0.0 | 3 | 3.1 |
| Asthma/reactive airway disease | 293 | 20.0 | 188 | 18.6 | 78 | 22.5 | 2 | 20.0 | 25 | 26.0 |
| Chronic liver disease | 0 | 0.0 | 0 | 0.0 | 0 | 0.0 | 0 | 0.0 | 0 | 0.0 |
| Congenital heart condition | 6 | 0.4 | 4 | 0.4 | 1 | 0.3 | 0 | 0.0 | 1 | 1.0 |
| Congenital lung condition | 3 | 0.2 | 2 | 0.2 | 0 | 0.0 | 1 | 10.0 | 0 | 0.0 |
| Down syndrome/chromosomal anomaly | 33 | 2.3 | 22 | 2.2 | 3 | 0.9 | 2 | 20.0 | 6 | 6.3 |
| Immunocompromised^c^ | 568 | 38.9 | 318 | 31.5 | 189 | 54.5 | 2 | 20.0 | 59 | 61.5 |
| Autoimmune disease | 68 | 4.7 | 27 | 2.7 | 40 | 11.5 | 0 | 0.0 | 1 | 1.0 |
| Sickle cell | 58 | 4.0 | 38 | 3.8 | 15 | 4.3 | 0 | 0.0 | 5 | 5.2 |
| Disability^d^ | 37 | 2.5 | 26 | 2.6 | 6 | 1.7 | 1 | 10.0 | 4 | 4.2 |
| Transplant (Bone marrow and organ) | 14 | 1.0 | 10 | 1.0 | 3 | 0.9 | 0 | 0.0 | 1 | 1.0 |

^a^ Unknown refers to either one of, or both, race and ethnicity are unknown.

^b^ Reported only top three discharge status here, remaining such as skilled nursing facility, home health organization, hospice, rehabilitation facility, and/or other are not listed.

^c^ Immunocompromised conditions included HIV/AIDS, malignancy, transplants, rheumatologic/other inflammatory conditions, primary immunodeficiency, CKD/ESRD, and other immune conditions.

^d^ Includes neurologic, neurodevelopmental, intellectual, physical, vision or hearing impairment.

AIDS: Acquired immunodeficiecy syndrome. CKD: Chronic kidney disease; ESRD: End stage renal disease; HIV: Human immunodeficiency virus; ICU: Intensive care unit; IMV: Invasive mechanical ventilation; PHD SR: Premier Healthcare Database Special Release; Q1: First quartile; Q3: Third quartile; SD: Standard deviation.

**Supplementary Table 3. Sensitivity Analysis ‒ HealthVerity RTIE database:** **Demographics, hospitalization characteristics, and comorbid conditions among children aged 0-11 years**

|  | **Overall study population  N=1962 (100%)** | | **Without ICU**  **or IMV  N=1481 (75.5%)** | | **With ICU, but without IMV  N=345 (17.6%)** | | **Without ICU, but with IMV  N=49 (2.5%)** | | **With ICU and IMV N=87 (4.4%)** | |
| --- | --- | --- | --- | --- | --- | --- | --- | --- | --- | --- |
| ***Age (years)*** |  |  |  |  |  |  |  |  |  |  |
| Median, Q1-Q3 | 2 | 0-7 | 2 | 0-7 | 3 | 1-8 | 2 | 0-6 | 3 | 1-6 |
| Mean, SD | 3.7 | 3.8 | 3.6 | 3.8 | 4.5 | 3.9 | 3.3 | 3.7 | 4.0 | 3.4 |
|  | ***N*** | ***%*** | ***N*** | ***%*** | ***N*** | ***%*** | ***N*** | ***%*** | ***N*** | ***%*** |
| ***Age group*** |  |  |  |  |  |  |  |  |  |  |
| 0-4 years | 1,246 | 63.5 | 969 | 65.4 | 190 | 55.1 | 35 | 71.4 | 52 | 59.8 |
| 5-11 years | 716 | 36.5 | 512 | 34.6 | 155 | 44.9 | 14 | 28.6 | 35 | 40.2 |
| ***Sex*** |  |  |  |  |  |  |  |  |  |  |
| Male | 1,090 | 55.6 | 822 | 55.5 | 190 | 55.1 | 26 | 53.1 | 52 | 59.8 |
| Female | 870 | 44.3 | 658 | 44.4 | 155 | 44.9 | 23 | 46.9 | 34 | 39.1 |
| Unknown | 2 | 0.1 | 1 | 0.1 | 0 | 0.0 | 0 | 0.0 | 1 | 1.1 |
| ***Insurance type*** |  |  |  |  |  |  |  |  |  |  |
| Commercial | 569 | 29.0 | 417 | 28.2 | 121 | 35.1 | 7 | 14.3 | 24 | 27.6 |
| Medicaid | 1,141 | 58.2 | 874 | 59.0 | 191 | 55.4 | 24 | 49.0 | 52 | 59.8 |
| Medicare | 23 | 1.2 | 15 | 1.0 | 8 | 2.3 | 0 | 0.0 | 0 | 0.0 |
| Unknown | 228 | 11.6 | 174 | 11.7 | 25 | 7.2 | 18 | 36.7 | 11 | 12.6 |
| Missing | 1 | 0.1 | 1 | 0.1 | 0 | 0.0 | 0 | 0.0 | 0 | 0.0 |
| ***Hospital Census region*** |  |  |  |  |  |  |  |  |  |  |
| Northeast | 305 | 15.5 | 233 | 15.7 | 46 | 13.3 | 8 | 16.3 | 18 | 20.7 |
| Midwest | 108 | 5.5 | 83 | 5.6 | 21 | 6.1 | 0 | 0.0 | 4 | 4.6 |
| South | 942 | 48.0 | 693 | 46.8 | 192 | 55.7 | 16 | 32.7 | 41 | 47.1 |
| West | 605 | 30.8 | 471 | 31.8 | 85 | 24.6 | 25 | 51.0 | 24 | 27.6 |
| Missing | 2 | 0.1 | 1 | 0.1 | 1 | 0.3 | 0 | 0.0 | 0 | 0.0 |
| ***Hospital population served*** |  |  |  |  |  |  |  |  |  |  |
| Urban | 1,694 | 86.3 | 1,268 | 85.6 | 315 | 91.3 | 38 | 77.6 | 73 | 83.9 |
| Rural | 236 | 12.0 | 184 | 12.4 | 30 | 8.7 | 8 | 16.3 | 14 | 16.1 |
| Unknown | 32 | 1.6 | 29 | 2.0 | 0 | 0.0 | 3 | 6.1 | 0 | 0.0 |
| ***COVID-19 diagnosis position*** |  |  |  |  |  |  |  |  |  |  |
| Primary | 1,024 | 52.2 | 777 | 52.5 | 188 | 54.5 | 20 | 40.8 | 39 | 44.8 |
| Secondary | 938 | 47.8 | 704 | 47.5 | 157 | 45.5 | 29 | 59.2 | 48 | 55.2 |
| ***Calendar period of admission*** |  |  |  |  |  |  |  |  |  |  |
| April 2020-April 2021 | 1,257 | 64.1 | 952 | 64.3 | 218 | 63.2 | 35 | 71.4 | 52 | 59.8 |
| May 2021-June 2021 | 93 | 4.7 | 75 | 5.1 | 15 | 4.3 | 0 | 0.0 | 3 | 3.4 |
| July 2021-September 2021 | 612 | 31.2 | 454 | 30.7 | 112 | 32.5 | 14 | 28.6 | 32 | 36.8 |
| ***Discharge status^b^*** |  |  |  |  |  |  |  |  |  |  |
| Home/self-care | 1,855 | 94.5 | 1,430 | 96.6 | 331 | 95.9 | 33 | 67.3 | 61 | 70.1 |
| Expired/died | 11 | 0.6 | 0 | 0.0 | 1 | 0.3 | 3 | 6.1 | 7 | 8.0 |
| Transferred to other facility | 31 | 1.6 | 26 | 1.8 | 1 | 0.3 | 3 | 6.1 | 1 | 1.1 |
| ***Comorbid conditions*** |  |  |  |  |  |  |  |  |  |  |
| Number of comorbidities: 0 | 1,070 | 54.5 | 885 | 59.8 | 145 | 42.0 | 20 | 40.8 | 20 | 23.0 |
| Number of comorbidities: 1-2 | 790 | 40.3 | 541 | 36.5 | 177 | 51.3 | 22 | 44.9 | 50 | 57.5 |
| Number of comorbidities: ≥3 | 102 | 5.2 | 55 | 3.7 | 23 | 6.7 | 7 | 14.3 | 17 | 19.5 |
| Median, Q1-Q3 | 0 | 0-0 | 0 | 0-0 | 0 | 0-0 | 0 | 0-2 | 0 | 0-2 |
| Mean, SD | 0.4 | 0.9 | 0.4 | 0.9 | 0.4 | 1.0 | 0.8 | 1.3 | 1.1 | 1.4 |
| ***Comorbid conditions*** |  |  |  |  |  |  |  |  |  |  |
| Diabetes | 41 | 2.1 | 22 | 1.5 | 17 | 4.9 | 1 | 2.0 | 1 | 1.1 |
| Obesity/overweight | 86 | 4.4 | 62 | 4.2 | 17 | 4.9 | 3 | 6.1 | 4 | 4.6 |
| Hypertension | 51 | 2.6 | 28 | 1.9 | 11 | 3.2 | 4 | 8.2 | 8 | 9.2 |
| CKD or ESRD | 108 | 5.5 | 53 | 3.6 | 26 | 7.5 | 9 | 18.4 | 20 | 23.0 |
| Neurological disease | 155 | 7.9 | 85 | 5.7 | 32 | 9.3 | 14 | 28.6 | 24 | 27.6 |
| Psychiatric disorders | 23 | 1.2 | 19 | 1.3 | 3 | 0.9 | 1 | 2.0 | 0 | 0.0 |
| Malignancy (solid or hematologic) | 74 | 3.8 | 59 | 4.0 | 10 | 2.9 | 1 | 2.0 | 4 | 4.6 |
| Asthma/reactive airway disease | 201 | 10.2 | 139 | 9.4 | 41 | 11.9 | 5 | 10.2 | 16 | 18.4 |
| Chronic liver disease | 0 | 0.0 | 0 | 0.0 | 0 | 0.0 | 0 | 0.0 | 0 | 0.0 |
| Congenital heart condition | 13 | 0.7 | 8 | 0.5 | 3 | 0.9 | 0 | 0.0 | 2 | 2.3 |
| Congenital lung condition | 3 | 0.2 | 1 | 0.1 | 0 | 0.0 | 0 | 0.0 | 2 | 2.3 |
| Down syndrome/chromosomal anomaly | 23 | 1.2 | 16 | 1.1 | 4 | 1.2 | 1 | 2.0 | 2 | 2.3 |
| Immunocompromised^c^ | 543 | 27.7 | 371 | 25.1 | 119 | 34.5 | 13 | 26.5 | 40 | 46.0 |
| Autoimmune disease | 40 | 2.0 | 25 | 1.7 | 14 | 4.1 | 0 | 0.0 | 1 | 1.1 |
| Sickle cell | 83 | 4.2 | 71 | 4.8 | 9 | 2.6 | 0 | 0.0 | 3 | 3.4 |
| Disability^d^ | 25 | 1.3 | 12 | 0.8 | 5 | 1.4 | 4 | 8.2 | 4 | 4.6 |
| Transplant (Bone marrow and organ) | 22 | 1.1 | 17 | 1.1 | 3 | 0.9 | 0 | 0.0 | 2 | 2.3 |

^a^ Unknown refers to either one of, or both, race and ethnicity are unknown.

^b^ Reported only top three discharge status here, remaining such as skilled nursing facility, home health organization, hospice, rehabilitation facility, and/or other are not listed.

^c^ Immunocompromised conditions included HIV/AIDS, malignancy, transplants, rheumatologic/other inflammatory conditions, primary immunodeficiency, CKD/ESRD, and other immune conditions.

^d^ Includes neurologic, neurodevelopmental, intellectual, physical, vision or hearing impairment.

CKD: Chronic kidney disease; ESRD: End stage renal disease; IBD: Inflammatory bowel disease; ICU: Intensive care unit; IMV: Invasive mechanical ventilation; Q1: First quartile; Q3: Third quartile; SD: Standard deviation.

**Supplementary Table 4. Sensitivity Analysis ‒ HealthVerity RTIE database: Demographics, hospitalization characteristics, and comorbid conditions among children aged 0-4 years**

|  | **Overall study population  N=1246 (100%)** | | **Without ICU**  **or IMV  N=969 (77.8%)** | | **With ICU, but without IMV  N=190 (15.2%)** | | **Without ICU, but with IMV  N=35 (2.8%)** | | **With ICU and IMV N=52 (4.2%)** | |
| --- | --- | --- | --- | --- | --- | --- | --- | --- | --- | --- |
| ***Age (years)*** |  |  |  |  |  |  |  |  |  |  |
| Median, Q1-Q3 | 1 | 0-2 | 0 | 0-2 | 1 | 0-2 | 0 | 0-3 | 1 | 0-3 |
| Mean, SD | 1.1 | 1.3 | 1.0 | 1.3 | 1.3 | 1.3 | 1.3 | 1.5 | 1.5 | 1.5 |
|  | ***N*** | ***%*** | ***N*** | ***%*** | ***N*** | ***%*** | ***N*** | ***%*** | ***N*** | ***%*** |
| ***Sex*** |  |  |  |  |  |  |  |  |  |  |
| Male | 681 | 54.7 | 538 | 55.5 | 94 | 49.5 | 20 | 57.1 | 29 | 55.8 |
| Female | 563 | 45.2 | 430 | 44.4 | 96 | 50.5 | 15 | 42.9 | 22 | 42.3 |
| Unknown | 2 | 0.2 | 1 | 0.1 | 0 | 0.0 | 0 | 0.0 | 1 | 1.9 |
| ***Insurance type*** |  |  |  |  |  |  |  |  |  |  |
| Commercial | 341 | 27.4 | 264 | 27.2 | 53 | 27.9 | 5 | 14.3 | 19 | 36.5 |
| Medicaid | 744 | 59.7 | 590 | 60.9 | 112 | 58.9 | 16 | 45.7 | 26 | 50.0 |
| Medicare | 18 | 1.4 | 12 | 1.2 | 6 | 3.2 | 0 | 0.0 | 0 | 0.0 |
| Unknown | 142 | 11.4 | 102 | 10.5 | 19 | 10.0 | 14 | 40.0 | 7 | 13.5 |
| Missing | 1 | 0.1 | 1 | 0.1 | 0 | 0.0 | 0 | 0.0 | 0 | 0.0 |
| ***Hospital Census region*** |  |  |  |  |  |  |  |  |  |  |
| Northeast | 186 | 14.9 | 149 | 15.4 | 22 | 11.6 | 5 | 14.3 | 10 | 19.2 |
| Midwest | 69 | 5.5 | 54 | 5.6 | 12 | 6.3 | 0 | 0.0 | 3 | 5.8 |
| South | 606 | 48.6 | 462 | 47.7 | 106 | 55.8 | 14 | 40.0 | 24 | 46.2 |
| West | 384 | 30.8 | 303 | 31.3 | 50 | 26.3 | 16 | 45.7 | 15 | 28.8 |
| Missing | 1 | 0.1 | 1 | 0.1 | 0 | 0.0 | 0 | 0.0 | 0 | 0.0 |
| ***Hospital population served*** |  |  |  |  |  |  |  |  |  |  |
| Urban | 1,081 | 86.8 | 832 | 85.9 | 173 | 91.1 | 28 | 80.0 | 48 | 92.3 |
| Rural | 142 | 11.4 | 116 | 12.0 | 17 | 8.9 | 5 | 14.3 | 4 | 7.7 |
| Unknown | 23 | 1.8 | 21 | 2.2 | 0 | 0.0 | 2 | 5.7 | 0 | 0.0 |
| ***COVID-19 diagnosis position*** |  |  |  |  |  |  |  |  |  |  |
| Primary | 681 | 54.7 | 539 | 55.6 | 110 | 57.9 | 11 | 31.4 | 21 | 40.4 |
| Secondary | 565 | 45.3 | 430 | 44.4 | 80 | 42.1 | 24 | 68.6 | 31 | 59.6 |
| ***Calendar period of admission*** |  |  |  |  |  |  |  |  |  |  |
| April 2020-April 2021 | 794 | 63.7 | 627 | 64.7 | 114 | 60.0 | 25 | 71.4 | 28 | 53.8 |
| May 2021-June 2021 | 57 | 4.6 | 47 | 4.9 | 10 | 5.3 | 0 | 0.0 | 0 | 0.0 |
| July 2021-September 2021 | 395 | 31.7 | 295 | 30.4 | 66 | 34.7 | 10 | 28.6 | 24 | 46.2 |
| ***Discharge status^b^*** |  |  |  |  |  |  |  |  |  |  |
| Home/self-care | 1,188 | 95.3 | 944 | 97.4 | 183 | 96.3 | 24 | 68.6 | 37 | 71.2 |
| Expired/died | 4 | 0.3 | 0 | 0.0 | 0 | 0.0 | 1 | 2.9 | 3 | 5.8 |
| Transferred to other facility | 14 | 1.1 | 11 | 1.1 | 0 | 0.0 | 2 | 5.7 | 1 | 1.9 |
| ***Comorbid conditions*** |  |  |  |  |  |  |  |  |  |  |
| Number of comorbidities: 0 | 853 | 68.5 | 709 | 73.2 | 110 | 57.9 | 18 | 51.4 | 16 | 30.1 |
| Number of comorbidities: 1-2 | 361 | 28.9 | 245 | 25.3 | 74 | 38.9 | 13 | 37.1 | 29 | 55.8 |
| Number of comorbidities: ≥3 | 32 | 2.6 | 15 | 1.6 | 6 | 3.2 | 4 | 11.4 | 7 | 13.5 |
| Median, Q1-Q3 | 0 | 0-0 | 0 | 0-0 | 0 | 0-0 | 0 | 0-0 | 0 | 0-2 |
| Mean, SD | 0.2 | 0.7 | 0.2 | 0.6 | 0.2 | 0.7 | 0.6 | 1.2 | 0.8 | 1.4 |
| ***Comorbid conditions^b^*** |  |  |  |  |  |  |  |  |  |  |
| Diabetes | 7 | 0.6 | 4 | 0.4 | 1 | 0.5 | 1 | 2.9 | 1 | 1.9 |
| Obesity/overweight | 12 | 1.0 | 8 | 0.8 | 2 | 1.1 | 2 | 5.7 | 0 | 0.0 |
| Hypertension | 31 | 2.5 | 17 | 1.8 | 5 | 2.6 | 3 | 8.6 | 6 | 11.5 |
| CKD or ESRD | 43 | 3.5 | 23 | 2.4 | 9 | 4.7 | 4 | 11.4 | 7 | 13.5 |
| Neurological disease | 71 | 5.7 | 31 | 3.2 | 13 | 6.8 | 10 | 28.6 | 17 | 32.7 |
| Psychiatric disorders | 3 | 0.2 | 0 | 0.0 | 2 | 1.1 | 1 | 2.9 | 0 | 0.0 |
| Malignancy (solid or hematologic) | 37 | 3.0 | 28 | 2.9 | 7 | 3.7 | 1 | 2.9 | 1 | 1.9 |
| Asthma/reactive airway disease | 55 | 4.4 | 32 | 3.3 | 16 | 8.4 | 2 | 5.7 | 5 | 9.6 |
| Chronic liver disease | 0 | 0.0 | 0 | 0.0 | 0 | 0.0 | 0 | 0.0 | 0 | 0.0 |
| Congenital heart condition | 10 | 0.8 | 5 | 0.5 | 3 | 1.6 | 0 | 0.0 | 2 | 3.8 |
| Congenital lung condition | 2 | 0.2 | 1 | 0.1 | 0 | 0.0 | 0 | 0.0 | 1 | 1.9 |
| Down syndrome/chromosomal anomaly | 13 | 1.0 | 11 | 1.1 | 1 | 0.5 | 0 | 0.0 | 1 | 1.9 |
| Immunocompromised^c^ | 261 | 20.9 | 188 | 19.4 | 49 | 25.8 | 6 | 17.1 | 18 | 34.6 |
| Autoimmune disease | 5 | 0.4 | 5 | 0.5 | 0 | 0.0 | 0 | 0.0 | 0 | 0.0 |
| Sickle cell | 43 | 3.5 | 39 | 4.0 | 3 | 1.6 | 0 | 0.0 | 1 | 1.9 |
| Disability^d^ | 9 | 0.7 | 4 | 0.4 | 1 | 0.5 | 1 | 2.9 | 3 | 5.8 |
| Transplant (Bone marrow and organ) | 8 | 0.6 | 7 | 0.7 | 1 | 0.5 | 0 | 0.0 | 0 | 0.0 |

^a^ Unknown refers to either one of, or both, race and ethnicity are unknown.

^b^ Reported only top three discharge status here, remaining such as skilled nursing facility, home health organization, hospice, rehabilitation facility, and/or other are not listed.

^c^ Includes immunocompromised conditions such as HIV/AIDS, malignancy, transplants, rheumatologic/other inflammatory conditions, primary immunodeficiency, CKD/ESRD, and other immune conditions.

^d^ Includes neurologic, neurodevelopmental, intellectual, physical, vision or hearing impairment.

CKD: Chronic kidney disease; ESRD: End stage renal disease; IBD: Inflammatory bowel disease; ICU: Intensive care unit; IMV: Invasive mechanical ventilation; Q1: First quartile; Q3: Third quartile; SD: Standard deviation.

**Supplementary Table 5. Sensitivity Analysis ‒ HealthVerity RTIE database: Demographics, hospitalization characteristics, and comorbid conditions among children aged 5-11 years**

|  | **Overall study population  N=716 (100%)** | | **Without ICU**  **or IMV  N=512 (71.5%)** | | **With ICU, but without IMV  N=155 (21.6%)** | | **Without ICU, but with IMV  N=14 (2.0%)** | | **With ICU and IMV N=35 (4.9%)** | |
| --- | --- | --- | --- | --- | --- | --- | --- | --- | --- | --- |
| ***Age (years)*** |  |  |  |  |  |  |  |  |  |  |
| Median, Q1-Q3 | 9 | 6-10 | 9 | 6-10 | 9 | 7-10 | 9 | 6-10 | 8 | 6-9 |
| Mean, SD | 8.3 | 2.1 | 8.3 | 2.1 | 8.4 | 2.1 | 8.4 | 2.2 | 7.6 | 2.0 |
|  | ***N*** | ***%*** | ***N*** | ***%*** | ***N*** | ***%*** | ***N*** | ***%*** | ***N*** | ***%*** |
| ***Sex*** |  |  |  |  |  |  |  |  |  |  |
| Male | 409 | 57.1 | 284 | 55.5 | 96 | 61.9 | 6 | 42.9 | 23 | 65.7 |
| Female | 307 | 42.9 | 228 | 44.5 | 59 | 38.1 | 8 | 57.1 | 12 | 34.3 |
| ***Insurance type*** |  |  |  |  |  |  |  |  |  |  |
| Commercial | 228 | 31.8 | 153 | 29.9 | 68 | 43.9 | 2 | 14.3 | 5 | 14.3 |
| Medicaid | 397 | 55.4 | 284 | 55.5 | 79 | 51.0 | 8 | 57.1 | 26 | 74.3 |
| Medicare | 5 | 0.7 | 3 | 0.6 | 2 | 1.3 | 0 | 0.0 | 0 | 0.0 |
| Unknown | 86 | 12.0 | 72 | 14.1 | 6 | 3.9 | 4 | 28.6 | 4 | 11.4 |
| ***Hospital Census region*** |  |  |  |  |  |  |  |  |  |  |
| Northeast | 119 | 16.6 | 84 | 16.4 | 24 | 15.5 | 3 | 21.4 | 8 | 22.9 |
| Midwest | 39 | 5.4 | 29 | 5.7 | 9 | 5.8 | 0 | 0.0 | 1 | 2.9 |
| South | 336 | 46.9 | 231 | 45.1 | 86 | 55.5 | 2 | 14.3 | 17 | 48.6 |
| West | 221 | 30.9 | 168 | 32.8 | 35 | 22.6 | 9 | 64.3 | 9 | 25.7 |
| Missing | 1 | 0.1 | 0 | 0.0 | 1 | 0.6 | 0 | 0.0 | 0 | 0.0 |
| ***Hospital population served*** |  |  |  |  |  |  |  |  |  |  |
| Urban | 613 | 85.6 | 436 | 85.2 | 142 | 91.6 | 10 | 71.4 | 25 | 71.4 |
| Rural | 94 | 13.1 | 68 | 13.3 | 13 | 8.4 | 3 | 21.4 | 10 | 28.6 |
| Unknown | 9 | 1.3 | 8 | 1.6 | 0 | 0.0 | 1 | 7.1 | 0 | 0.0 |
| ***COVID-19 diagnosis position*** |  |  |  |  |  |  |  |  |  |  |
| Primary | 343 | 47.9 | 238 | 46.5 | 78 | 50.3 | 9 | 64.3 | 18 | 51.4 |
| Secondary | 373 | 52.1 | 274 | 53.5 | 77 | 49.7 | 5 | 35.7 | 17 | 48.6 |
| ***Calendar period of admission*** |  |  |  |  |  |  |  |  |  |  |
| April 2020-April 2021 | 463 | 64.7 | 325 | 63.5 | 104 | 67.1 | 10 | 71.4 | 24 | 68.6 |
| May 2021-June 2021 | 36 | 5.0 | 28 | 5.5 | 5 | 3.2 | 0 | 0.0 | 3 | 8.6 |
| July 2021-September 2021 | 217 | 30.3 | 159 | 31.1 | 46 | 29.7 | 4 | 28.6 | 8 | 22.9 |
| ***Discharge status^b^*** |  |  |  |  |  |  |  |  |  |  |
| Home/self-care | 667 | 93.2 | 486 | 94.9 | 148 | 95.5 | 9 | 64.3 | 24 | 68.6 |
| Expired/died | 7 | 1.0 | 0 | 0.0 | 1 | 0.6 | 2 | 14.3 | 4 | 11.4 |
| Transferred to other facility | 17 | 2.4 | 15 | 2.9 | 1 | 0.6 | 1 | 7.1 | 0 | 0.0 |
| ***Comorbid conditions*** |  |  |  |  |  |  |  |  |  |  |
| Number of comorbidities: 0 | 217 | 30.3 | 176 | 34.4 | 35 | 22.6 | 2 | 14.3 | 4 | 11.4 |
| Number of comorbidities: 1-2 | 429 | 59.9 | 296 | 57.8 | 103 | 66.5 | 9 | 64.3 | 21 | 60.0 |
| Number of comorbidities: ≥3 | 70 | 9.8 | 40 | 7.8 | 17 | 11.0 | 3 | 21.4 | 10 | 28.6 |
| Median, Q1-Q3 | 0 | 0-2 | 0 | 0-2 | 0 | 0-2 | 2 | 0-2 | 2 | 0-3 |
| Mean, SD | 0.7 | 1.2 | 0.7 | 1.1 | 0.7 | 1.1 | 1.4 | 1.4 | 1.7 | 1.2 |
| ***Comorbid conditions*** |  |  |  |  |  |  |  |  |  |  |
| Diabetes | 34 | 4.7 | 18 | 3.5 | 16 | 10.3 | 0 | 0.0 | 0 | 0.0 |
| Obesity/overweight | 74 | 10.3 | 54 | 10.5 | 15 | 9.7 | 1 | 7.1 | 4 | 11.4 |
| Hypertension | 20 | 2.8 | 11 | 2.1 | 6 | 3.9 | 1 | 7.1 | 2 | 5.7 |
| CKD or ESRD | 65 | 9.1 | 30 | 5.9 | 17 | 11.0 | 5 | 35.7 | 13 | 37.1 |
| Neurological disease | 84 | 11.7 | 54 | 10.5 | 19 | 12.3 | 4 | 28.6 | 7 | 20.0 |
| Psychiatric disorders | 20 | 2.8 | 19 | 3.7 | 1 | 0.6 | 0 | 0.0 | 0 | 0.0 |
| Malignancy (solid or hematologic) | 37 | 5.2 | 31 | 6.1 | 3 | 1.9 | 0 | 0.0 | 3 | 8.6 |
| Asthma/reactive airway disease | 146 | 20.4 | 107 | 20.9 | 25 | 16.1 | 3 | 21.4 | 11 | 31.4 |
| Chronic liver disease | 0 | 0.0 | 0 | 0.0 | 0 | 0.0 | 0 | 0.0 | 0 | 0.0 |
| Congenital heart condition | 3 | 0.4 | 3 | 0.6 | 0 | 0.0 | 0 | 0.0 | 0 | 0.0 |
| Congenital lung condition | 1 | 0.1 | 0 | 0.0 | 0 | 0.0 | 0 | 0.0 | 1 | 2.9 |
| Down syndrome/chromosomal anomaly | 10 | 1.4 | 5 | 1.0 | 3 | 1.9 | 1 | 7.1 | 1 | 2.9 |
| Immunocompromised^c^ | 282 | 39.4 | 183 | 35.7 | 70 | 45.2 | 7 | 50.0 | 22 | 62.9 |
| Autoimmune disease | 35 | 4.9 | 20 | 3.9 | 14 | 9.0 | 0 | 0.0 | 1 | 2.9 |
| Sickle cell | 40 | 5.6 | 32 | 6.3 | 6 | 3.9 | 0 | 0.0 | 2 | 5.7 |
| Disability^d^ | 16 | 2.2 | 8 | 1.6 | 4 | 2.6 | 3 | 21.4 | 1 | 2.9 |
| Transplant (Bone marrow and organ) | 14 | 2.0 | 10 | 2.0 | 2 | 1.3 | 0 | 0.0 | 2 | 5.7 |

^a^ Unknown refers to either one of, or both, race and ethnicity are unknown.

^b^ Reported only top three discharge status here, remaining such as skilled nursing facility, home health organization, hospice, rehabilitation facility, and/or other are not listed.

^c^ Includes immunocompromised conditions such as HIV/AIDS, malignancy, transplants, rheumatologic/other inflammatory conditions, primary immunodeficiency, CKD/ESRD, and other immune conditions.

^d^ Includes neurologic, neurodevelopmental, intellectual, physical, vision or hearing impairment.

CKD: Chronic kidney disease; ESRD: End stage renal disease; IBD: Inflammatory bowel disease; ICU: Intensive care unit; IMV: Invasive mechanical ventilation; Q1: First quartile; Q3: Third quartile; SD: Standard deviation.

**Supplementary Table 6. Sensitivity Analysis ‒ HealthVerity RTIE database: Health outcomes and costs among children aged 0-11 years**

|  | **Index hospitalizations** | **Hospital LOS, days** | | **In-hospital mortality** | **Hospital costs^a^** | | **Hospital charges** | |
| --- | --- | --- | --- | --- | --- | --- | --- | --- |
|  | N  (%) | Median  (Q1-Q3) | Mean  (SD) | N  (%) | Median  (Q1-Q3) | Mean  (SD) | Median  (Q1-Q3) | Mean  (SD) |
| **All inpatient admissions** | 1962  (100) | 3  (2-5) | 5.2  (6.1) | 11  (0.6) | $10,286  ($5827-$16,855) | $15,893  ($19,745) | $25,166  ($13,083-$61,267) | $62,953  ($125,353) |
| ***Without ICU or IMV*** | 1481  (75.5) | 3  (2-5) | 4.1  (3.8) | 0  (0) | $9071  ($5402-$14,806) | $12,580  ($14,561) | $20,429  ($10,822-$41,327) | $37,043  ($53,771) |
| ***With ICU, but without IMV*** | 345  (17.6) | 4  (3-7) | 6.1  (5.6) | 1  (0.3) | $14,388  ($8607-$23,833) | $21,218  ($22,499) | $55,938  ($28,003-$110,232) | $83,519  ($82,916) |
| **Without ICU, but with IMV** | 49  (2.5) | 12  (5-22) | 15.9  (14.7) | 3  (6.1) | $26,124  ($17,371-$32,314) | $29,476  ($25,358) | $161,410  ($77,976-$450,823) | $330,944  ($449,163) |
| ***With ICU and IMV*** | 87  (4.4) | 9  (5-17) | 13.1  (14.3) | 7  (8.0) | $35,528  ($15,995-$64,719) | $45,364  ($40,755) | $182,488  ($78,868-$368,008) | $271,531  ($265,056) |
|  | **Readmissions** | **Hospital LOS, days** | | **In-hospital mortality** | **Hospital costs^a^** | | **Hospital charges** | |
| **Index hospitalization status** | N  (%)^b^ | Median  (Q1-Q3) | Mean  (SD) | N  (%) | Median  (Q1-Q3) | Mean  (SD) | Median  (Q1-Q3) | Mean  (SD) |
| **All inpatient admissions** | 51  (2.6) | 4  (3-6) | 5.1  (3.5) | 1  (2.0) | $10,671  ($6100-$20,079) | $14,868  ($12,639) | $31,644  ($13,690-$66,768) | $54,611  ($67,048) |
| ***Without ICU or IMV*** | 44  (3.0) | 4  (3-5.5) | 4.7  (2.6) | 1  (2.3) | $8,859  ($4,963-$18,223) | $13,590  ($11,514) | $30,383  ($13,294-$60,563) | $44,375  ($40,174) |
| ***With ICU, but without IMV*** | 5  (1.5) | 9  (3-11) | 9.2  (7.6) | 0  (0) | $26,514  ($6761-$46,910) | $26,728  ($20,075) | $94,171  ($16,173-$136,679) | $135,240  ($170,229) |
| **Without ICU, but with IMV** | 1  (2.0) | 4  - | 4  - | 0  (0) | $11,256  - | $11,256  - | $53,411  - | $53,411  - |
| ***With ICU and IMV*** | 1  (1.2) | 4  - | 4  - | 0  (0) | $8,463  - | $8,463  - | $103,054  - | $103,054  - |

^a^ Among patients with validated costs >$0 (N=1,578 for index hospitalizations; N=25 for readmissions).

^b^ Percentage among those with COVID-19-associated index hospitalizations, overall and stratified by the 4 COVID-19 disease progression states.

ICU: Intenstive care unit; IMV: Invasive mechanical ventilation; LOS: Length of stay; Q1: First quartile; Q3: Third quartile; SD: Standard deviation

**Supplementary Table 7 Sensitivity Analysis ‒ HealthVerity RTIE database: Health outcomes and costs among children aged 0-4 years**

|  | **Index hospitalizations** | **Hospital LOS, days** | | **In-hospital mortality** | **Hospital costs^a^** | | **Hospital charges** | |
| --- | --- | --- | --- | --- | --- | --- | --- | --- |
|  | N  (%) | Median  (Q1-Q3) | Mean  (SD) | N  (%) | Median  (Q1-Q3) | Mean  (SD) | Median  (Q1-Q3) | Mean  (SD) |
| **All inpatient admissions** | 1246  (100) | 3  (2-5) | 5.0  (6.5) | 7  (0.6) | $9201  ($5383-$15,046) | $13,654  ($17,627) | $21,203  ($11,055-$44,543) | $52,971  ($114,257) |
| ***Without ICU or IMV*** | 969  (77.8) | 3  (2-4) | 3.9  (4.0) | 1  (0.1) | $8125  ($5117-$13,225) | $10,923  ($12,136) | $17,798  ($9502-$32,032) | $31,216  ($52,069) |
| ***With ICU, but without IMV*** | 190  (15.2) | 4  (3-6) | 5.8  (5.9) | 0  (0) | $12,385  ($7410-$16,650) | $16,604  ($17,426) | $38,555  ($20,993-$76,100) | $64,194  ($74,198) |
| **Without ICU, but with IMV** | 35  (2.8) | 8  (5-22) | 15.4  (13.5) | 2  (5.7) | $26,124  ($14,638-$32,314) | $28,512  ($26,328) | $158,468  ($50,853-$450,823) | $289,789  ($333,029) |
| ***With ICU and IMV*** | 52  (4.2) | 8  (3.5-18.5) | 14.0  (17.3) | 4  (7.7) | $35,336  ($9564-$67,479) | $46,699  ($43,984) | $135,810  ($65,254-$318,117) | $257,955  ($286,526) |
|  | **Readmissions** | **Hospital LOS, days** | | **In-hospital mortality** | **Hospital costs^a^** | | **Hospital charges** | |
| **Index hospitalization status** | N  (%)^b^ | Median  (Q1-Q3) | Mean  (SD) | N  (%) | Median  (Q1-Q3) | Mean  (SD) | Median  (Q1-Q3) | Mean  (SD) |
| **All inpatient admissions** | 31  (2.5) | 4  (2-6) | 5.1  (4.3) | 0  (0) | $7048  ($6100-$14,686) | $12,306  ($10,802) | $30,559  ($12,533-$63,980) | $56,188  ($81,884) |
| ***Without ICU or IMV*** | 25  (2.6) | 4  (2-5) | 4.5  (3.1) | 0  (0) | $6955  ($5037-$14,686) | $12,015  ($11,610) | $24,451  ($12,533-$46,292) | $40,133  ($44,153) |
| ***With ICU, but without IMV*** | 4  (2.1) | 7  (2.5-16) | 9.3  (8.8) | 0  (0) | $16,638  ($6761-$26,514) | $16,638  ($13,967) | $76,426  ($10,826-$280,189) | $145,508  ($194,768) |
| **Without ICU, but with IMV** | 1  (2.9) | 4  - | 4  - | 0  (0) | $11,256  - | $11,256  - | $53,411  - | $53,411  - |
| ***With ICU and IMV*** | 1  (1.9) | 4  - | 4  - | 0  (0) | $8,463  - | $8,463  - | $103,054  - | $103,054  - |

^a^ Among patients with validated costs >$0 (N=991 for index hospitalizations; N=17 for readmissions).

^b^ Percentage among those with COVID-19-associated index hospitalizations, overall and stratified by the 4 COVID-19 disease progression states.

ICU: Intenstive care unit; IMV: Invasive mechanical ventilation; LOS: Length of stay; Q1: First quartile; Q3: Third quartile; SD: Standard deviation

**Supplementary Table 8. Sensitivity Analysis ‒ HealthVerity RTIE database: Health outcomes and costs among children aged 5-11 years**

|  | **Index hospitalizations** | **Hospital LOS, days** | | **In-hospital mortality** | **Hospital costs^a^** | | **Hospital charges** | |
| --- | --- | --- | --- | --- | --- | --- | --- | --- |
|  | N  (%) | Median  (Q1-Q3) | Mean  (SD) | N  (%) | Median  (Q1-Q3) | Mean  (SD) | Median  (Q1-Q3) | Mean  (SD) |
| **All inpatient admissions** | 716  (100) | 4  (3-7) | 5.5  (5.3) | 9  (1.3) | $12,351  ($7241-$22,034) | $19,674  ($22,393) | $41,054  ($18,812-$94,583) | $80,325  ($141,029) |
| ***Without ICU or IMV*** | 512  (71.5) | 4  (2-5) | 4.5  (3.3) | 2  (0.4) | $11,070  ($6796-$17,446) | $15,662  ($17,843) | $28,168  ($14,939-$57,854) | $48,071  ($55,244) |
| ***With ICU, but without IMV*** | 155  (21.6) | 6  (3-8) | 6.6  (5.1) | 1  (0.6) | $16,648  ($9975-$35,506) | $26,449  ($26,232) | $86,508  ($43,887-$146,805) | $107,206  ($87,035) |
| **Without ICU, but with IMV** | 14  (2.0) | 13.5  (5-23) | 17.1  (18.0) | 2  (14.3) | $26,124  ($17,780-$39,264) | $31,753  ($23,956) | $246,995  ($116,087-$459,386) | $433,830  ($662,484) |
| ***With ICU and IMV*** | 35  (4.9) | 11  (5-16) | 11.7  (8.1) | 4  (11.4) | $37,700  ($17,966-$57,390) | $43,521  ($36,513) | $266,661  ($114,334-$394,822) | $291,701  ($232,069) |
|  | **Readmissions** | **Hospital LOS, days** | | **In-hospital mortality** | **Hospital costs^a^** | | **Hospital charges** | |
| **Index hospitalization status** | N  (%)^b^ | Median  (Q1-Q3) | Mean  (SD) | N  (%) | Median  (Q1-Q3) | Mean  (SD) | Median  (Q1-Q3) | Mean  (SD) |
| **All inpatient admissions** | 20  (2.8) | 4  (4-6) | 5.1  (2.0) | 1  (5.0) | $18,223  ($7217-$30,539) | $20,313  ($15,203) | $42,728  ($27,173-$69,957) | $49,230  ($34,283) |
| ***Without ICU or IMV*** | 19  (3.7) | 4  (4-6) | 4.9  (1.8) | 1  (5.3) | $16,368  ($3764-$26,025) | $16,513  ($11,615) | $39,905  ($26,935-$66,767) | $46,865  ($33,504) |
| ***With ICU, but without IMV*** | 1  (0.7) | 9  - | 9  - | 0  (0) | $46,910  - | $46,910  - | $94,171  - | $94,171  - |
| **Without ICU, but with IMV** | 0  (0) | - | - | 0  (0) | - | - | - | - |
| ***With ICU and IMV*** | 0  (0) | - | - | 0  (0) | - | - | - | - |

^a^ Among patients with validated costs >$0 (N=587 for index hospitalizations; N=8 for readmissions).

^b^ Percentage among those with COVID-19-associated index hospitalizations, overall and stratified by the 4 COVID-19 disease progression states.

ICU: Intenstive care unit; IMV: Invasive mechanical ventilation; LOS: Length of stay; Q1: First quartile; Q3: Third quartile; SD: Standard deviation

**Supplementary Table 9. Sensitivity Analysis ‒ HealthVerity RTIE database: Premier database: Index hospitalizations by calendar period among children aged 0-11 years**

|  | **Index hospitalizations: Apr 2020-Apr 2021** | **Hospital LOS, days** | | **In-hospital mortality** | **Hospital costs^a^** | | **Hospital charges** | |
| --- | --- | --- | --- | --- | --- | --- | --- | --- |
|  | N  (%) | Median  (Q1-Q3) | Mean  (SD) | N  (%) | Median  (Q1-Q3) | Mean  (SD) | Median  (Q1-Q3) | Mean  (SD) |
| **All inpatient admissions** | 1257  (100) | 3  (3-5) | 5.3  (6.5) | 14  (1.1) | $10,613  ($6203-$17,558) | $16,380  ($20,103) | $27,196  ($13,895-$64,795) | $68,613  ($138,629) |
| ***Without ICU or IMV*** | 952  (75.7) | 3  (2-5) | 4.3  (4.1) | 3  (0.3) | $9119  ($5478-$15,174) | $12,679  ($13,860) | $21,072  ($11,751-$44,708) | $41,327  ($61,210) |
| ***With ICU, but without IMV*** | 218  (17.3) | 4  (3-7) | 6.1  (5.0) | 1  (0.5) | $14,702  ($8911-$25,139) | $22,344  ($23,796) | $55,002  ($28,162-$111,020) | $84,679  ($82,716) |
| **Without ICU, but with IMV** | 35  (2.8) | 8  (4-21) | 15.0  (15.0) | 4  (11.4) | $26,124  ($17,371-$34,148) | $30,561  ($26,534) | $168,291  ($77,976-$493,138) | $372,533  ($506,282) |
| ***With ICU and IMV*** | 52  (4.1) | 10  (4.5-18) | 14.8  (17.3) | 6  (11.5) | $42,256  ($18,523-$65,095) | $50,389  ($42,881) | $251,187  ($80,949-$386,570) | $296,261  (268,703) |
|  | **Index hospitalizations: May-Jun 2021** | **Hospital LOS, days** | | **In-hospital mortality** | **Hospital costs^a^** | | **Hospital charges** | |
|  | N  (%)^b^ | Median  (Q1-Q3) | Mean  (SD) | N  (%) | Median  (Q1-Q3) | Mean  (SD) | Median  (Q1-Q3) | Mean  (SD) |
| **All inpatient admissions** | 93  (100) | 3  (2-4) | 3.8  (2.8) | 0  (0) | $8725  ($5509-$14,233) | $12,509  ($11,757) | $21,998  ($11,327-$55,117) | $40,047  ($49,833) |
| ***Without ICU or IMV*** | 75  (80.6) | 3  (2-4) | 3.3  (1.9) | 0  (0) | $8344  ($5107-$12,177) | $10,238  ($8911) | $19,349  ($10,769-$31,669) | $27,954  ($27,181) |
| ***With ICU, but without IMV*** | 15  (16.1) | 4  (2-8) | 5.5  (4.7) | 0  (0) | $17,921  ($6876-$30,641) | $19,946  ($15,036) | $67,569  ($21,998-$99,356) | $71,461  ($60,706) |
| **Without ICU, but with IMV** | 0  (0) | - | - | - | - | - | - | - |
| ***With ICU and IMV*** | 3  (3.2) | 6  (5-13) | 8.0  (4.4) | 0  (0) | $16,779  ($6848-$57,390) | $27,006  ($26,778) | $127,566  ($95,578-$332,766) | $185,303  ($128,704) |
|  | **Index hospitalizations: Jul-Sep 2021** | **Hospital LOS, days** | | **In-hospital mortality** | **Hospital costs^a^** | | **Hospital charges** | |
|  | N  (%) | Median  (Q1-Q3) | Mean  (SD) | N  (%) | Median  (Q1-Q3) | Mean  (SD) | Median  (Q1-Q3) | Mean  (SD) |
| **All inpatient admissions** | 612  (100) | 3  (2-5) | 5.1  (5.5) | 2  (0.3) | $9718  ($5271-$15,995) | $15,272  ($20,011) | $23,651  ($11,209-$53,630) | $54,809  ($101,690) |
| ***Without ICU or IMV*** | 454  (74.2) | 3  (2-5) | 4  (3.3) | 0  (0) | $9000  ($4535-$14,059) | $12,808  ($17,111) | $19,231  ($9069-$34,285) | $29,563  ($36,766) |
| ***With ICU, but without IMV*** | 112  (18.3) | 4  (3-7) | 6.4  (6.6) | 0  (0) | $13,240  ($8741-$16,902) | $18,409  ($19,940) | $55,339  ($29,622-$103,604) | $82,875  ($86,274) |
| **Without ICU, but with IMV** | 14  (2.3) | 17  (6-26) | 18.1  (14.2) | 0  (0) | $22,269  ($8658-$27,841) | $23,868  ($18,987) | $149,930  ($46,554-$254,184) | $226,970  ($241,315) |
| ***With ICU and IMV*** | 32  (5.2) | 8  (4.5-16.5) | 10.7  (8.0) | 2  (6.3) | $24,485  ($6301-$60,027) | $37,816  ($32,707) | $136,220  ($56,896-$259,100) | $239,429  ($268,774) |

^a^ Among patients with validated costs >$0.

^b^ Percentage among those with COVID-19-associated index hospitalizations, overall and stratified by the 4 COVID-19 disease progression states.

ICU: Intenstive care unit; IMV: Invasive mechanical ventilation; LOS: Length of stay; Q1: First quartile; Q3: Third quartile; SD: Standard deviation

**Supplementary Table 10. Sensitivity Analysis ‒ HealthVerity RTIE database: Readmission outcomes by calendar period among children aged 0-11 years**

| **Index hospitalization status** | **Index hospitalizations: Apr 2020-Apr 2021** | **Hospital LOS, days** | | **Hospital costs^a^** | | **Hospital charges** | |
| --- | --- | --- | --- | --- | --- | --- | --- |
|  | N  (%) | Median  (Q1-Q3) | Mean  (SD) | Median  (Q1-Q3) | Mean  (SD) | Median  (Q1-Q3) | Mean  (SD) |
| **All inpatient admissions**  **N=1257** | 37  (2.9) | 4  (3-5) | 5.0  (3.7) | $9567  ($6100-$20,079) | $14,874  ($13,068) | $36,866  ($12,898-$63,980) | $57,286  ($74,992) |
| ***Without ICU or IMV  N=952*** | 31  (3.3) | 4  (3-5) | 4.3  (2.2) | $7048  ($5037-$16,368) | $13,372  ($11,900) | $30,559  ($12,533-$55,325) | $42,035  ($40,220) |
| ***With ICU, but without IMV  N=218*** | 4  (1.8) | 10  (5.5-16) | 10.8  (7.9) | $26,514  ($6,762-$46,910) | $26,728  ($20,075) | $115,425  ($49,825-$280,189) | $165,007  ($180,915) |
| **Without ICU, but with IMV**  ***N=35*** | 1  (2.9) | 4  - | 4  - | $11,256  - | $11,256  - | $53,411  - | $53,411  - |
| ***With ICU and IMV***  ***N=52*** | 1  (1.9) | 4  - | 4  - | $8,463  - | $8,463  - | $103,054  - | $103,054  - |
|  | **Index hospitalizations: May 2021-Jun 2021** | **Hospital LOS, days** | | **Hospital costs^a^** | | **Hospital charges** | |
| **Index hospitalization status** | N  (%) | Median  (Q1-Q3) | Mean  (SD) | Median  (Q1-Q3) | Mean  (SD) | Median  (Q1-Q3) | Mean  (SD) |
| **All inpatient admissions**  **N=93** | 0  (0) | - | - | - | - | - | - |
| ***Without ICU or IMV  N=75*** | 0  (0) | - | - | - | - | - | - |
| ***With ICU, but without IMV  N=15*** | 0  (0) | - | - | - | - | - | - |
| **Without ICU, but with IMV**  ***N=0*** | 0  (0) | - | - | - | - | - | - |
| ***With ICU and IMV***  **N=3** | 0  (0) | - | - | - | - | - | - |
|  | **Index hospitalizations: Jul 2021-Sep 2021** | **Hospital LOS, days** | | **Hospital costs^a^** | | **Hospital charges** | |
| **Index hospitalization status** | N  (%)^b^ | Median  (Q1-Q3) | Mean  (SD) | Median  (Q1-Q3) | Mean  (SD) | Median  (Q1-Q3) | Mean  (SD) |
| **All inpatient admissions**  **N=612** | 14  (2.3) | 4  (3-6) | 5.4  (3.3) | $14,686  ($3764-$26,025) | $14,824  ($11,131) | $26,684  ($16,173-$73,146) | $47,542  ($40,528) |
| ***Without ICU or IMV  N=454*** | 13  (2.9) | 4  (3-6) | 5.6  (3.3) | $14,686  ($3764-$26,025) | $14,824  ($11,131) | $30,208  ($16,561-$73,146) | $49,955  ($41,123) |
| ***With ICU, but without IMV  N=112*** | 1  (0.9) | 3  - | 3  - | -  - | -  - | $16,173  - | $16,173  - |
| **Without ICU, but with IMV**  ***N=14*** | 0  (0) | - | - | - | - | - | - |
| ***With ICU and IMV***  **N=32** | 0  (0) | - | - | - | - | - | - |

^a^ Among patients with validated costs >$0.

^b^ Percentage among those with COVID-19-associated index hospitalizations, overall and stratified by the 4 COVID-19 disease progression states.

ICU: Intenstive care unit; IMV: Invasive mechanical ventilation; LOS: Length of stay; Q1: First quartile; Q3: Third quartile; SD: Standard deviation

**Supplementary Figure 1. Sensitivity Analysis ‒ HealthVerity RTIE database: Frequency of index hospitalizations by calendar month among children aged 0-11 years**

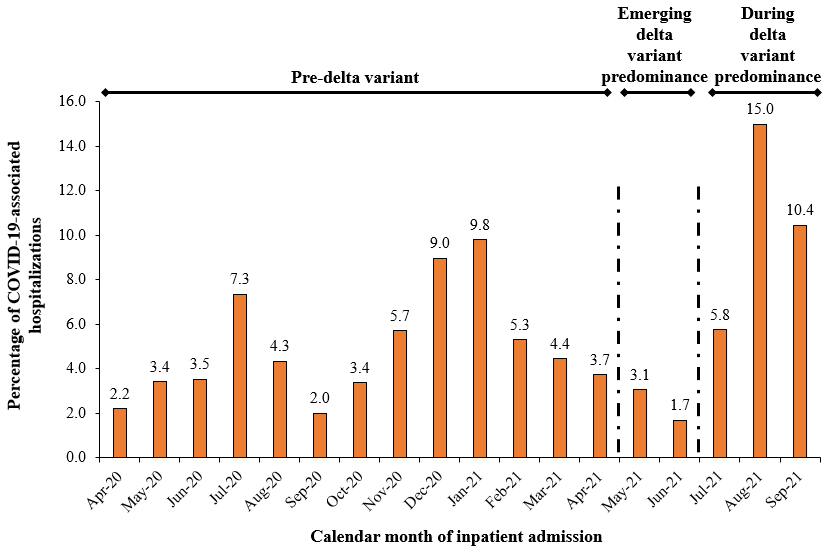
